# The natural history of TB disease-a synthesis of data to quantify progression and regression across the spectrum

**DOI:** 10.1101/2021.09.13.21263499

**Authors:** Alexandra S Richards, Bianca Sossen, Jon C Emery, Katherine C Horton, Torben Heinsohn, Beatrice Frascella, Federica Balzarini, Aurea Oradini-Alacreu, Brit Hacker, Anna Odone, Nicky McCreesh, Alison D Grant, Katharina Kranzer, Frank Cobelens, Hanif Esmail, Rein MGJ Houben

## Abstract

**Background:** Prevalence surveys have found a substantial burden of subclinical (asymptomatic but infectious) TB, from which individuals can progress, regress or even persist in a chronic disease state. We aimed to quantify these pathways across the spectrum of TB disease.

**Methods:** We created a deterministic framework of TB disease with progression and regression between three states of pulmonary TB disease: minimal (non-infectious), subclinical, and clinical (symptomatic and infectious) disease. We estimated ranges for each parameter by considering all data from a systematic review in a Bayesian framework, enabling quantitative estimation of TB disease pathways.

**Findings:** Twenty-two studies contributed data from 5942 individuals. Results suggested that, after five years, 39.5%(95% uncertainty interval, UI, 31.4%-47.3%) of individuals with prevalent subclinical disease at baseline had recovered after regressing to minimal disease and 18.4%(95%UI, 13.7%-24%) had died from TB, leaving 14%(95%UI, 10.1%- 18.6%) individuals still with infectious disease at five years, and the remainder with minimal disease at risk of further progression. Over the course of five years 50.2% (95%UI, 41.1%-59%) of the subclinical cohort never developed symptoms. For those with clinical disease at baseline, 45.9%(95%UI, 38.9%-52.1%) and 19.8%(95%UI, 15.1%-25.3%) had died or recovered from TB respectively, with the remainder in, or undulating between, the three disease states. The ten-year mortality of people with untreated prevalent infectious disease was 38%.

**Interpretation:** Our results show that for people with subclinical disease, classic clinical disease is neither inevitable nor an irreversible outcome. As such, reliance on symptom-based screening means a large proportion of people with infectious disease may never be detected.

**Funding:** TB Modelling and Analysis Consortium and European Research Council

**Research in Context:** *Evidence before this study:* In recent years the existence of a spectrum of TB disease has been re-accepted. The classic paradigm of disease is one active state of symptomatic presentation with bacteriologically positive sputum, now referred to as clinical disease. Within the spectrum, a subclinical phase (where people do not report symptoms but have bacteriologically positive sputum) has been widely accepted, due to prevalence surveys using chest radiography screening in addition to symptom screening. On average these prevalence surveys have found around 50% of people with prevalent infectious TB had subclinical disease. There is also another state of minimal disease, or non-infectious disease, that is the earliest point on the disease spectrum after progression from infection. The likelihood or speed of natural progression, regression, or persistence of individuals across this spectrum remains unknown. As a consequence, the ability to accurately predict the impact of interventions has been limited. As individuals with bacteriologically-positive TB now receive treatment, contemporary data to inform the required transitions is highly limited. However, a large number of cohorts of patients were described in the pre-chemotherapy era. Until now, these data have not been synthesised to inform parameters to describe the natural history of TB disease.

*Added value of this study:* We synthesised data from historical and contemporary literature to explore the expected trajectories of individuals across the spectrum of TB disease. We considered a cohort of people with prevalent bacteriologically positive disease, with a 50/50 split of people with subclinical and clinical disease at baseline. We found that within five years, 29.6% of people recover from TB, defined as no chance of progressing to active disease without reinfection. However, we also find that 13.7% are still spending time infectious at the end of the five years. Our estimates for 10 year mortality and duration of symptoms before treatment aligned with the known and accepted values. We also show that regression from subclinical disease results in a large reservoir of people with minimal disease, from which they can permanently recover, but can also progress again to subclinical disease. The undulating pathways that lead to regression and progression mean that 50.2% (41.1%-59%) of individuals with prevalent subclinical disease do not experience symptoms over the course of five years. This shows that clinical disease is neither a rapid, nor inevitable outcome of subclinical disease.

*Implications of the available evidence:* With these data-driven estimates of parameters, informed projections of the relative value of addressing minimal, subclinical, or clinical disease can now be provided. Given the known reservoir of prevalent subclinical disease and its contribution to transmission, efforts to diagnose and treat people with “earlier” stages of TB are likely to have a larger impact than strategies targeting clinical disease, particularly on individuals who never would have progressed to clinical disease.

## Introduction

Despite effective treatment regimens being discovered in the 1950s, tuberculosis (TB) is still a major cause of morbidity and mortality globally. In 2019, there were an estimated 10 million people who fell ill with TB, and 1.4 million people died from TB.^1^

The current paradigm of TB disease assumes that there is a single state of active disease, with only progression to active disease from infection.^1^ In reality people can move in both directions across a spectrum of disease.^2,3^ After initial infection, individuals transitioning to pulmonary disease have been shown to progress through a state of minimal disease, where pathological changes due to *Mycobacterium tuberculosis* (*Mtb*) are visible on imaging techniques such as chest radiography (CXR) or computed tomography (CT), but individuals are not infectious (bacteriologically negative sputum).^4–6^ Further progression leads to infectious disease (bacteriologically positive), within which there is a distinction between clinical and subclinical disease where individuals with subclinical disease do not report symptoms, but individuals with clinical disease report a prolonged cough or seek treatment due to their symptoms.^2,4,7^ A recent review of national TB prevalence surveys found that around 50% of people with prevalent infectious disease have subclinical disease, and therefore will not be diagnosed by policies that rely on reported symptoms.^8^

While it is likely that not all individuals with minimal or subclinical disease will progress to clinical disease, the range or relative significance of alternative disease pathways is effectively unknown.^5,9,10^ Efforts to quantify these pathways have remained limited by the absence of directly applicable parameter estimates.^11–13^ A comprehensive review of literature has shown that many data sources exist, both historical and contemporary, which observed cohorts transitioning across the spectrum of disease.^14^ However, no single study provides the overview of all trajectories across the different states, and with studies having varying durations, follow-up structures, and approaches to define and report disease states, the resulting heterogeneity complicates a simple comprehensive analysis.

Here we use a Bayesian framework to synthesise all available data to inform estimates of the natural rate of transition between minimal, subclinical, and clinical TB disease. We use these rates to simulate disease pathways in individuals, which we categorise to compare the frequency of different disease pathways in the population.

## Methods

### Data

The systematic review collected data that describe untreated cohorts at a minimum of two time points. Each time point, the cohort disease state was reported with a combination of CXR, bacteriology, and symptoms, with all studies required to directly report bacteriology or use standards set by the National Tuberculosis Association (NTA) that include bacteriology in the definitions.^6,14^ The first time point described the state of a baseline group, and the second (and further) time points described the states of a subgroup after a recorded time. To enable synthesised analysis, two study types were included. In cumulative follow-up studies, individuals were closely followed and cumulatively recorded whether they had transitioned to a new state, either with a single, or multiple consecutive reporting points. After transitioning, individuals were excluded from follow-up. In cross-sectional follow-up studies, individuals were followed up at the single reported time point. Only their final state was recorded, without knowledge of any additional transitions that occurred before the study end. For inclusion in the analysis, a study needed to report on at least one cohort that transitioned between states, and included individuals needed to have, as a minimum, a CXR with signs interpreted as TB activity to fit in the minimal disease category. Detail on inclusion and exclusion criteria are included in the appendix (section S4).

Minimal disease was defined as bacteriologically negative, regardless of symptoms, based on observation from numerous studies that did not report differing progression rates and the poor specificity of symptoms in bacteriologically positive TB (see appendix section S3).^9,15^ We adjusted the cohort size for people starting with minimal disease based on using tuberculin skin tests (TST) as a proxy for radiography changes that were truly caused by *Mtb* infection (see appendix, section S5.4). Whilst the systematic review collected data on whether a CXR was considered active or inactive, this distinction has not been carried forward here with a single grouping of minimal for both (see appendix, section S5.4).

For classification of outcomes, we assumed that when symptoms were only reported at enrolment, the symptom status persisted over the course of the study. Where symptom status was unknown at both time points, we classified people with bacteriologically positive disease as “infectious” as they could not be differentiated by symptoms to split between subclinical and clinical disease. If a paper referenced the NTA standards and used disease terminology of arrested, quiescent, or active from these standards, we have interpreted these to mean minimal, subclinical, and clinical respectively (see appendix sections S1 and S4).^6^

Both recovery from minimal disease and death from clinical disease were estimated through the calibration without data from the systematic review.^14^ We assumed no knowledge on recovery, providing a uniform prior from 0 to 12 per year (representing no recovery to recovery of everyone within a month) as seen in table 1. For death from clinical disease, the prior was taken from the estimated rate based on empirical data for mortality from “open” TB, which has a similar definition to clinical disease (see appendix section S2)^6,16,17^

**Table 1:** Posterior parameters calculated from the fit. The parameters are presented as annual rates, on the timescale of a year, so a parameter value of one means that, in the absence of any other competing parameters, the mean duration of disease in the initial state is one year. Priors with a uniform distribution are presented with the minimum and maximum value, priors with a normal distribution are presented with the median value and standard deviation

| Parameter | Prior | Posterior estimate<br>(95% uncertainty<br>interval) | Source/comments |
| --- | --- | --- | --- |
| recovery from<br>minimal | Uniform:<br>0-12/yr | 0.19<br>(0.15, 0.24) | Model calibration |
| minimal to<br>subclinical | Uniform:<br>0-12/yr | 0.25<br>(0.22, 0.29) | Data synthesis/model<br>calibration |
| subclinical to<br>minimal | Uniform:<br>0-12/yr | 1.52<br>(1.19, 1.94) | Data synthesis/model<br>calibration |
| subclinical to<br>clinical | Uniform:<br>0-12/yr | 0.69<br>(0.54, 0.89) | Data synthesis/model<br>calibration |
| clinical to<br>subclinical | Uniform:<br>0-12/yr | 0.57<br>(0.46, 0.71) | Data synthesis/model<br>calibration |
| death from clinical | Normal:<br>$\mu = 0.389/\text{yr}$<br>$\sigma = 0.028$ | 0.32<br>(0.27, 0.38) | Ragonnet et al/model<br>calibration.<sup>16</sup> |
| duration of<br>infectious disease | Normal:<br>$\mu = 2 \text{ yrs}$<br>$\sigma = 0.5$ | 0.99<br>(0.9, 1.11) | From NTI study/model<br>calibration.<sup>18</sup> See appendix,<br>section S5.3 |
| prevalence ratio<br>subclinical:clinical | Normal:<br>$\mu = 1 \text{ yr}$<br>$\sigma = 0.25$ | 1.3<br>(1, 1.68) | Frascella et al, Onozaki et<br>al/model calibration.<sup>19</sup> |
| prevalence ratio<br>minimal:infectious | Normal:<br>$\mu = 2.5\text{yrs}$<br>$\sigma = 0.5$ | 3.93<br>(3.44, 4.52) | Mungai et al/model<br>calibration.<sup>20</sup> |

Three further data points were included to inform the fit. Firstly, the median duration of infectious disease was included as a prior in the model assuming a normal distribution with a mean of 2 years and a standard deviation of 6 months (see appendix, section S5.3).^18^ The other two data points relate to the distribution of disease at steady state. Firstly, the ratio of subclinical to clinical disease was included with a normally distributed prior with a mean of 1 and standard deviation of 0.25 to match a recent review of prevalence surveys (see appendix section S5.3).^19^ Secondly, the ratio of minimal to infectious disease was included with a normally distributed prior with a mean of 2.5 and wide standard deviation of 0.5 (see appendix section S5.3).^20^ All these priors are outlined in table 1. The equations used to calculate and fit to each of these priors are derived in appendix sections S5.2 and S5.3.

### Data synthesis

To bring the data together we created a deterministic framework of TB disease, including the potential to move between the three disease states, as well as recovery from minimal disease and death from clinical TB disease (figure 1, top row).

**Figure 1:**
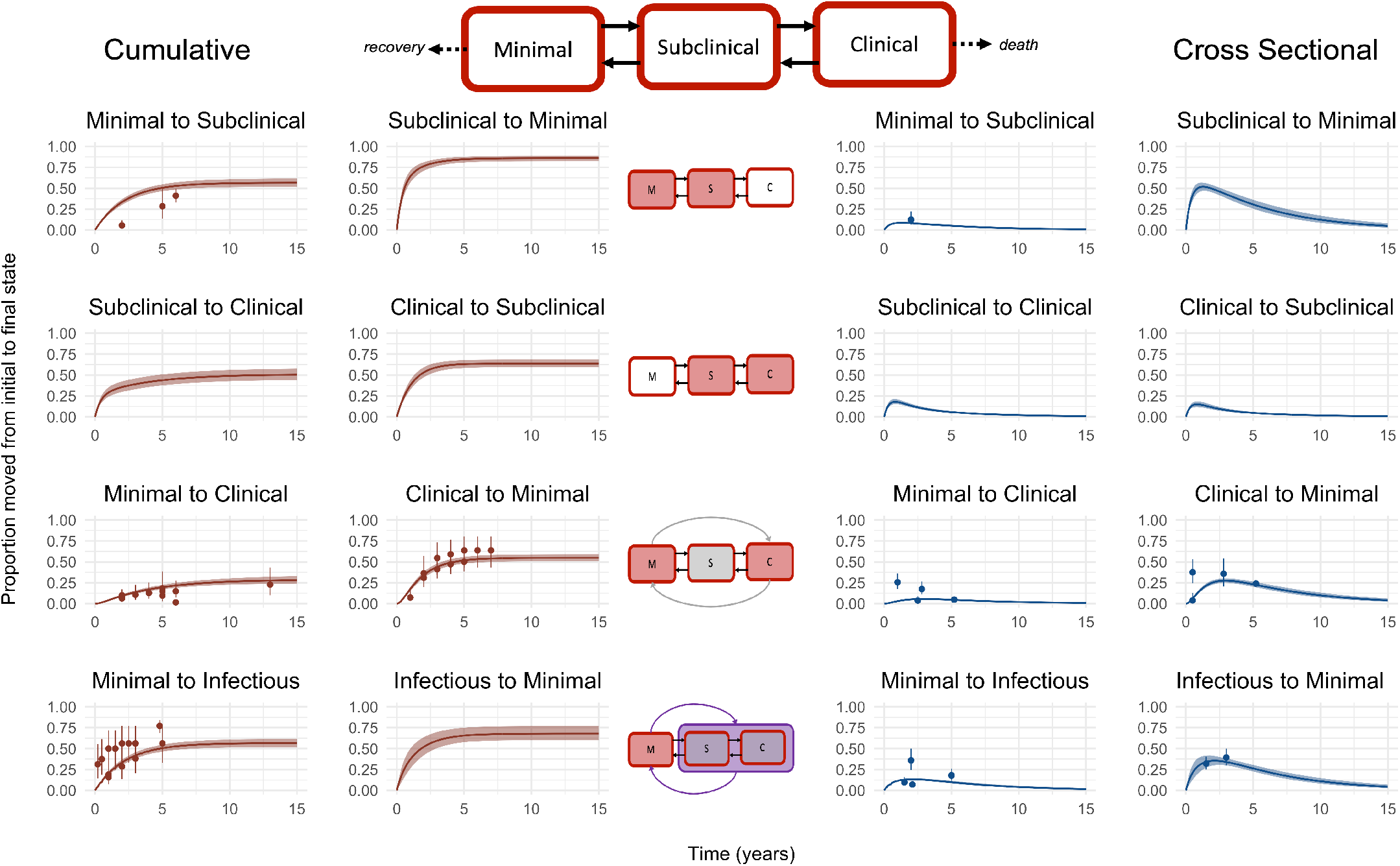
The model structure and the results of the fitting process compared to the data provided. The first row shows the model structure, with the solid lines representing parameters fitted by the model and the dotted line representing fixed parameters. Each small figure shows a unique combination of data transition and data type. The two left hand columns are the fits for the cumulative data, and the two right hand columns are the fits for the cross-sectional data. The rows show progression and regression between; 1) minimal and subclinical, 2) subclinical and clinical, 3) minimal and clinical, 4) minimal and infectious. The middle column is a visual description of the transition being fitted on each row. The dots in each graph are the point values provided from each study, with the error bars representing the weighting of that point value as provided in the fit (see appendix section S5.1). The solid line represents the median trajectory of that transition with the cloud covering 95% of the simulated trajectories

The transition rates were estimated by fitting to the data in a Bayesian framework. All data were considered simultaneously and a binomial distribution was chosen for the likelihood which allowed weighting by cohort size. Data points from cumulative studies were down-weighted so that the multiple data points of a cohort contributed as a single study (see appendix section S5.1).

We sampled the posterior values using a sequential Markov chain Monte-Carlo method (MCMC). An initial burn-in phase was used to find an optimal acceptance level, of between 25 and 35%, which was achieved by adapting the proposal distributions in both shape and scale. This was then discarded leaving chains with 10,000 iterations, which were visually inspected for convergence. The posterior parameter estimates came directly from the output of these chains, including the distribution, median, and 95% uncertainty intervals.

### TB disease pathways

To quantify the different pathways through disease, we applied the parameter values from the Bayesian fitting to a cohort model that tracked individuals through their TB disease history. Once recovered or treated, individuals exited the model. As we were interested in the natural trajectory of an existing disease episode, we did not include re-infection. Each cohort tracked 10,000 people over 10 years and this was repeated 1,000 times with the parameters re-sampled at each repeat to capture uncertainty. We considered three cohort types; subclinical cohorts where all individuals initially had subclinical disease, clinical cohorts where all individuals initially had clinical disease, and mixed cohorts where half the individuals had subclinical disease and the other half had clinical disease.^8^

We ran the cohorts with and without the possibility of diagnosis and treatment. When included, treatment was implemented as a rate of 0.7 per year defining the chance of being diagnosed and successfully treated while symptomatic, as a simple approximation of a 70% case detection rate in a care system reliant on self-reported symptoms to initiate the care pathway.

We categorised the different pathways of disease observed over 12 month intervals. Where an individual received treatment, died, or recovered during those 12 months, they were classified as such. Individuals not classified as one of those outcomes could either have a static disease state or were classified as undulating (i.e. moving between two or more disease states). The cohort model reports disease state monthly, and if fewer than nine of the 12 months were spent in a single state, or an individual transitioned between states three or more times, the disease pathway was classified as undulating. Otherwise, nine months or more in a single disease state, and fewer than three transitions is classified as a static state, of the dominant state during that interval. See appendix section S6 for examples of these trajectories.

We report two durations of disease, one for infectious disease (subclinical and clinical), and one for all TB disease (minimal, subclinical, and clinical). The median duration of disease was calculated as the first point after the start of the simulation that fewer than 50% of the original cohort are present in one of the relevant states. We also recorded the number of months an individual spent with clinical disease before treatment or death, as well as throughout their disease episode, regardless of outcome.

Cumulative mortality from infectious TB disease in the absence of treatment was recorded at 10 years to allow comparison with existing estimates based on historical data.^21^

### Sensitivity analyses

To test the robustness of the data synthesis results, we explored the impact of removing data provided from each study one at a time. In addition, the priors for mortality and the median duration of infectious disease were varied. For studies where symptoms were only provided in the start state of minimal, we re-ran the analysis with the transition for those studies to infectious rather than inferring a final state based on the initial symptoms. Sensitivities on the further analyses were also conducted; testing the parameter selection in the cohort model, introducing treatment at different case detection rates, and varying the thresholds for the definition of undulating disease.

All analyses were conducted in R version 4.0.3, using RStudio version 1.4.1103 and the Bayesian calibration was performed in LibBi version 1.4.5_3, using RBi version 0.10.3 and rbi.helpers version 0.3.2 as the interface.^22,23^

## Results

### Data synthesis

Twenty-two studies were included from the systematic review, providing 54 data points, describing 5942 people of whom 1034 transitioned between disease states. These cohorts were followed for intervals between 1923 and 2004, with studies conducted in North America (6), Europe (7), Asia (7), and one each from South America and Africa. In total there were 5 data points from minimal to subclinical, 14 data points from minimal to clinical, 15 data points from clinical to minimal, 18 data points from minimal to infectious, and 2 data points from infectious to minimal. Figure 1 shows the data points, including the relative weight of each data point, as indicated by the error bars. The best fit and uncertainty intervals to the data are shown by the lines and shaded area respectively in each plot. These data and the fitting are described in more detail in the appendix, sections S4 and S5.

Table 1 gives the median posterior parameter estimate for each model transition, with the 95% uncertainty interval. Uncertainty intervals for the parameters reflect the restricted parameter space when considering all the data simultaneously. Regression parameters were consistently higher than progression parameters.

### TB disease pathways

Figure 2 shows the relative proportions of each trajectory each year over the five years for simulated individuals with prevalent subclinical (figure 2A) and clinical (figure 2B) disease, as described in the methods. At five years, the proportion of people who die from TB is higher in the simulated cohort that starts with clinical disease (45.9%, 95% uncertainty interval, UI, 38.9%-52.1% vs 18.4%, 95%UI 13.7%-24%). The proportion of people in the minimal and recovered states is higher in the cohort starting with subclinical disease than it is in a cohort starting with clinical disease. Two-thirds of individuals with subclinical TB had regressed to minimal or recovered after five years (67.4%, 95%UI, 53.9%-81.6%), compared to 40.3%, 95%UI, 31.6%-50.1% of individuals with clinical TB. At the end of five years, regardless of the initial state, approximately one in eight of the cohort remain in subclinical, clinical, or undulating in or out of those states (14%, 95%UI, 10.2%-18.5% for subclinical compared to 13.7%, 95%UI, 10.1%-18.2% for clinical).

**Figure 2:**
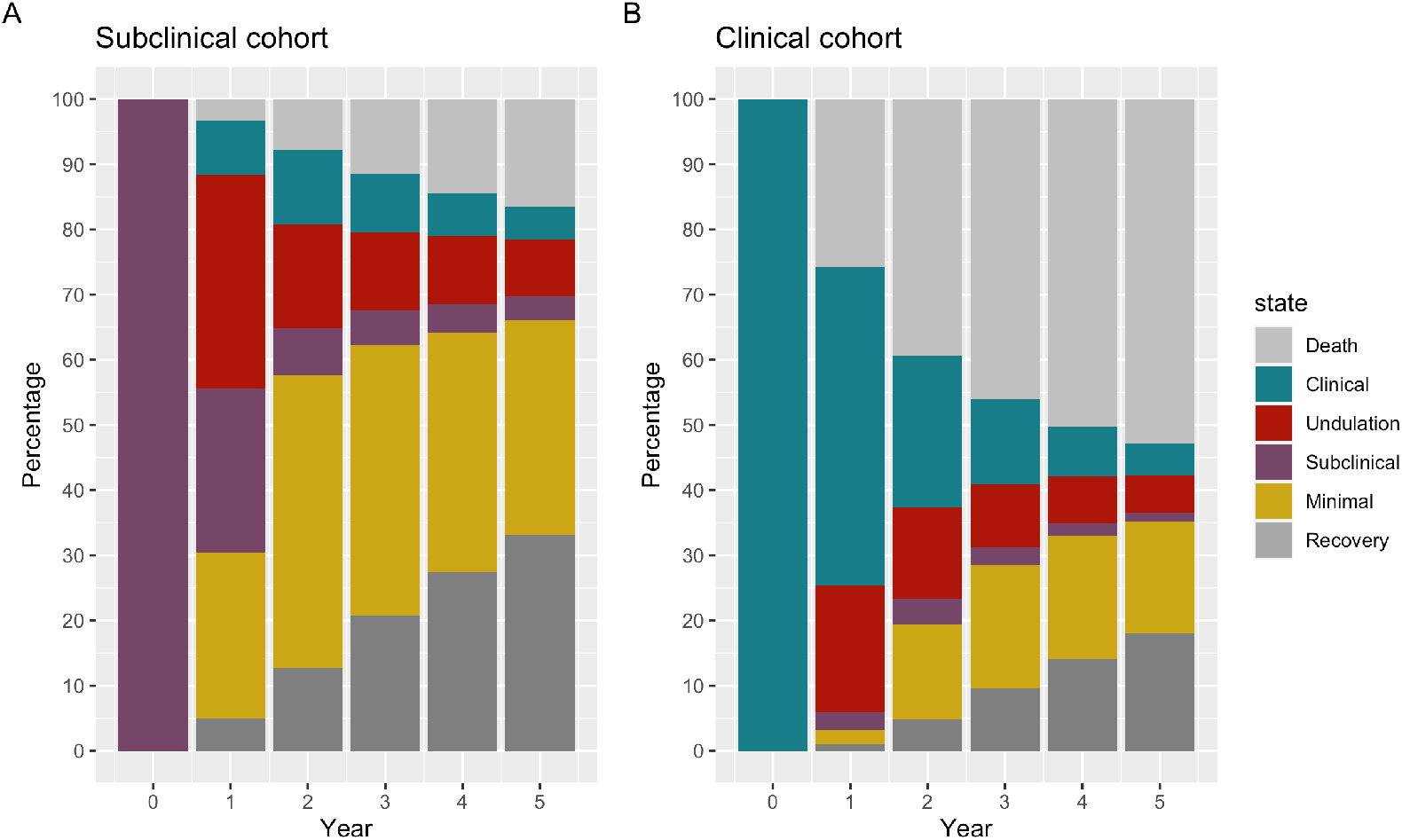
Trajectories of disease over time given different cohort starts

Figure 3 shows the start and end states of a mixed subclinical and clinical cohort over five years. It highlights the patterns seen in Figure 2 with fewer people remaining with minimal disease at five years from the clinical half of the cohort, but similar numbers of people with clinical or subclinical disease from each half of the cohort. Death is twice as likely from the clinical half of cohort while recovery is twice as likely in the subclinical half of the cohort.

**Figure 3:**
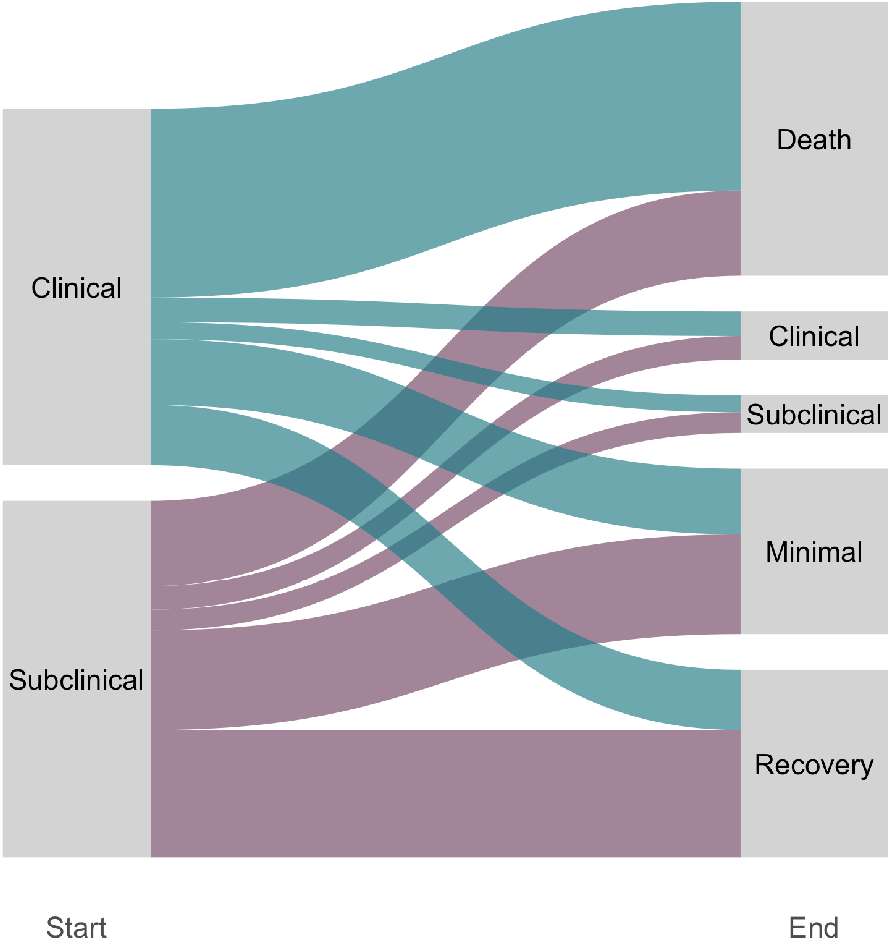
Final state after five years of people starting in subclinical and clinical disease

Looking at how many simulated individuals with subclinical disease at baseline never develop clinical disease, of those who completely recover within five years, 83.2% (95%UI, 78.9%-87.1%) never developed symptoms. This drops to 54.2% (95%UI, 45.8%-62.3%) in those with minimal disease at the end of five years, and further still to 25.9% (95%UI, 16%- 38.7%) for those with subclinical disease at the end of five years. In total, in a cohort of individuals with subclinical disease at baseline, we estimate that 50.2% (95%UI, 41.1%- 59%) would never develop symptoms.

In the absence of treatment, for a mixed cohort of half subclinical and half clinical at baseline, the median duration of infectious disease (subclinical and clinical) was 12 months (95%UI 10 - 15). If diagnosis and treatment were included, the median infectious period dropped to 8 months. However, we estimate the median duration of all TB disease, including the minimal state from which individuals can progress to infectious disease, to be 45 months without treatment and 35 with treatment. Illustrative figures can be found in the appendix, section S8.

In a simulated cohort with treatment available the duration of symptoms before death, regression to subclinical disease, or treatment varied between individuals from 1 months to 64 months, with a median of 6 months. For more on this distribution, see appendix section S7.

In a mixed cohort 37.7% (95%UI, 31.2%-45.2%) of the cohort died from TB within 10 years. A cohort of people with clinical disease at baseline had a higher proportion die from TB than one with subclinical disease at baseline (51.2% (95%UI, 43.6%-58.2%) vs 24.2% (95%UI, 18.1%-31.9%)), as seen in figure 3.

### Sensitivity analyses

Where changes in median parameter values were observed in the sensitivity analyses, these did not transfer through to the other key output metrics. In-depth comparisons of the sensitivity analyses can be found in the appendix, section S9.

## Discussion

### Summary of findings

We synthesized available data from a systematic review of untreated cohorts of TB disease to parameterise progression and regression between minimal, subclinical, and clinical TB disease. With these parameters we quantified the pathways of individuals across the spectrum. Our results show that non-linear disease trajectories are common, and there is a high rate of natural regression from subclinical disease. This is demonstrated by the large proportion of individuals at five years who have regressed to minimal disease or fully recovered. Although the risk of death from clinical disease is high, of those who do not die, the majority have also regressed to minimal disease or have recovered over the course of five years. Of those who still have infectious disease after five years, regardless of starting point, over half undulate between states rather than remaining with a single state of disease long term. Where symptom screening is used to detect people with infectious disease, many people may not be offered timely treatment due to intermittent or absent symptoms, meaning either they progress to more severe disease or unknowingly contribute to further transmission of TB.

### Interpretations

Of the four parameters estimated through fitting to the data, we found the regression rate from subclinical to minimal greatly exceed the other rates. While a high rate of recovery from subclinical to minimal could suggest that the majority of infectious TB disease would resolve without intervention, the relative sizes of the states as well as the high mortality rate from clinical disease likely counteract this trend, as can be seen in figure 3. For example, given many more individuals become infected with *Mtb* than develop infectious disease, it is reasonable to assume that the population with minimal disease is much larger than those with clinical disease which could mean the absolute number of individuals with potential to progress towards infectious disease will exceed those regressing.^1^ A rate of 1.52 per year would translate to a mean duration of 8 months with subclinical disease before regressing to minimal disease when not considering the transition to clinical disease, so although the rate is much higher than the other parameters, the duration it represents is not insignificant. The probability of recurrence of disease in individuals who have regressed to minimal disease is also higher than the probability of fully recovering which creates a loop of undulation, where an individual progresses and regresses in and out of infectious disease. As such, the benefits of wider treatment may be greater than has previously been assumed. For example, the rapid decline of prevalent infectious TB in China could be explained in part by the extensive treatment of individuals based on CXR alone in addition to those with a bacteriological diagnosis, which would have reduced the reservoir of minimal disease, as the combined rate of treatment and recovery exceeded that of progression from minimal.^24^

Each of the parameters are presented with a 95% uncertainty interval, some of which are narrower than others. Notably, the uncertainty interval around the value for subclinical to minimal is the largest of the main parameters. This may reflect the lack of data to directly fit this transition. However, when viewed in figure 1, these wide intervals do not translate to overwhelmingly wide bands, illustrating how the simultaneous consideration of all data restrict the potential parameter values that provide a reasonable fit to all the data points. In practice, progression and regression will be more variable between individuals and between populations, driven by variations in e.g. HIV status, diabetes, malnutrition, and gender.^28^ The cohorts represented in this analysis mostly comprised HIV-negative individuals, and the prevalence of other variables was unknown (e.g. malnutrition). While it is possible that these factors affect all or a subset of parameters, our results are based on a range of populations, times, and geography, and thereby provide an improvement over the limited data currently supporting estimates of TB progression and regression parameters.^16,29^

The parameters were estimated by fitting to the data collected from the systematic review alongside a prior of a two year infectious disease duration, and the ratios of subclinical to clinical disease and minimal to infectious disease at a steady state.^8,14,18–20^ The posterior estimate for infectious disease duration is at the lower end of the expected range set in the prior, however the data that informed the prior estimate had variation cannot be represented with a point value.^18,21^ We used previously reported values on TB mortality from smear positive disease as the prior for TB mortality from clinical disease.^16^ Although our point estimate was slightly lower than the provided prior, the 10-year mortality for people with untreated prevalent infectious TB was comparable to the accepted value of 40%.^16,21^ We also extracted duration of symptoms before treatment. Systematic reviews of self-reported symptom duration usually state between one and three months of symptoms prior to treatment, whereas we found a median of six months.^30^ While our results find a longer duration, it is likely that self-reported symptoms are more under-than over-estimated.

We cannot directly compare our parameter estimates with current models, as no other models have split the disease spectrum into three states. Ku et al used the proportion subclinical in prevalence surveys to divide total duration of disease, and did not consider undulation between states, however, our estimate of a median of 6 months falls within the range of symptom duration reported.^31^ The WHO technical appendix includes regression from “active” disease for those who self-cure or die before treatment as a single parameter with no further consideration of a spectrum of disease.^32^ Salvatore et al have represented disease as progression and regression along a single continuum of disease burden, defined as a composite of bacteriology, pathology, and symptoms.^11^ The potential rates of progression and regression were wide and overlapped with our data driven estimates.^11^ A recent systematic review from Menzies et al on progression only considered a single disease state.^29^ Some of these studies split the active disease state by bacteriological load (smear positive or smear negative, whilst still bacteriologically positive), however we have instead focused on bacteriological positivity alone, in line with the current reporting framework.^1^

We reported undulating disease based on a fixed threshold of nine months, which is a subjective choice. While a shift in threshold would change the proportion qualified as “undulating”, the underlying movement between states will remain the same (see appendix, sections S6 and S9.2.3).

An important finding is the large proportion of people with subclinical disease who may never develop symptoms, i.e. clinical disease. Although our results suggest many regress towards minimal disease, or even recover completely, this does not mitigate the time these people spend with infectious subclinical disease or the non-infectious period where the *Mtb* infection remains active. As we show, in a population without treatment, almost half of those who had subclinical disease at baseline, still have TB disease five years later.

### Limitations

Despite the extensive literature review, few data points could directly inform parameters. By including data on transitions between minimal and clinical, and between minimal and infectious, we were able to restrict the likely parameter space. For example, the cumulative minimal to infectious data provide a lower bound for the minimal to subclinical transition. While the chosen model structure will drive some of the results, limitations in the available data prohibit a more complicated model structure. In addition our three-state linear model structure is in line with historical and recent conceptualisations of the spectrum of TB disease.^2,3,6,8,13^

Both our data and simulations start from prevalent disease (minimal, subclinical, and clinical) without knowledge of previous disease trajectory and a single rate of transition for all. As such the parameters represent a mix of both recent and more distal *Mtb* infections, where some individuals are rapidly progressing, as well as individuals who are undulating, or on their way to recovery. However this is a reflection of current prevalent TB states in a population, as found in prevalence surveys.^8,9^ Prevalent disease is the immediate driver of TB morbidity, mortality, and transmission, and as such the population that TB policies look to address.

## Conclusions

We estimate that only half of all people with subclinical TB disease will progress to clinical disease. As such we show a flaw in the assumption that targeting clinical disease will enable care for all individuals suffering from TB disease, or interrupt transmission from infectious disease. Our work also highlights an important question; where should the threshold be set for TB disease that requires treatment. While the current threshold of infectious disease can relatively easily be confirmed, minimal (i.e. bacteriologically-negative) disease is an important reservoir of potential future transmission in the population and has a substantial risk of progression to more advanced disease. To comprehensively interrupt current and future transmission, we need to expand our interventions to those which can detect and treat subclinical and even minimal disease if possible. These may have substantial individual and population benefits which with these parameter estimates, we can now more reliably quantify.^10,13,33^

## Data Availability

All data is available in the supplementary materials and references

## Acknowledgements

ASR, JCE, KCH and RMGJH received funding from the European Research Council (ERC) under the European Union’s Horizon 2020 programme (Starting Grant Action Number 757699). KCH is supported by the UK FCDO (Leaving no-one behind: transforming gendered pathways to health for TB). This research has been partially funded by UK aid from the UK government (to KCH); however the views expressed do not necessarily reflect the UK government’s official policies.

HE, BS, received funding from the TB Modelling and Analysis Consortium (TB MAC)

HE received funding from the Medical Research Council (Grant ref: MR/V00476X/1)

NM is funded by the Wellcome Trust (218261/Z/19/Z)

FC received no funding for this work. KK received no funding for this work.

FB, BF, AOA, AO received no funding for this work.

## References

1 WHO. Global tuberculosis report 2020. 2020 https://www.who.int/publications-detail-redirect/9789240013131 (accessed 10 Jun2021).

2 Barry CE, Boshoff HI, Dartois V, Dick T, Ehrt S, Flynn J et al. The spectrum of latent tuberculosis: Rethinking the biology and intervention strategies. Nat Rev Microbiol 2009; 7: 845–855.

3 Gothi GD. NATURAL HISTORY OF TUBERCULOSIS. Indian Journal of Tuberculosis 1977; 25.

4 Drain PK, Bajema KL, Dowdy D, Dheda K, Naidoo K, Schumacher SG et al. Incipient and subclinical tuberculosis: A clinical review of early stages and progression of infection. Clinical Microbiology Reviews 2018; 31: 24.

5 Esmail H, Lai RP, Lesosky M, Wilkinson KA, Graham CM, Coussens AK et al. Characterization of progressive HIV-associated tuberculosis using 2-deoxy-2-[18F]fluoro-d-glucose positron emission and computed tomography. Nat Med 2016; 22: 1090–3.

6 Association NT. Diagnostic standards and classification of tuberculosis. 1940 ed. New York, N.Y., 1940http://hdl.handle.net/2027/coo.31924089435949.

7 Pai M, Behr MA, Dowdy D, Dheda K, Divangahi M, Boehme CC et al. Tuberculosis. Nat Rev Dis Primers 2016; 2: 16076.

8 Frascella B, Richards AS, Sossen B, Emery JC, Odone A, Law I et al. Subclinical tuberculosis disease - a review and analysis of prevalence surveys to inform definitions, burden, associations and screening methodology. Clin Infect Dis 2020. doi:10.1093/cid/ciaa1402.

9 Okada K, Onozaki I, Yamada N, Yoshiyama T, Miura T, Saint S et al. Epidemiological impact of mass tuberculosis screening: A 2 year follow-up after a national prevalence survey. 2012; NA.

10 Houben RMGJ, Esmail H, Emery JC, Joslyn LR, McQuaid CF, Menzies NA et al. Spotting the old foe—revisiting the case definition for TB. The Lancet Respiratory Medicine 2019; 7: 199–201.

11 Salvatore PP, Proano A, Kendall EA, Gilman RH, Dowdy DW. Linking individual natural history to population outcomes in tuberculosis. J Infect Dis 2017. doi:10.1093/infdis/jix555.

12 Arinaminpathy N, Dowdy D. Understanding the incremental value of novel diagnostic tests for tuberculosis. Nature 2015; 528: S60–7.

13 Dowdy DW, Basu S, Andrews JR. Is passive diagnosis enough? Am J Respir Crit Care Med 2013; 187: 543–551.

14 Sossen B. Systematic review. 2021.

15 Organisation. WH. Rapid communication on systematic screening for tuberculosis. World Health Organisation, 2020https://extranet.who.int/iris/restricted/bitstream/handle/10665/337372/9789240016552-eng.pdf (accessed 7 Dec2020).

16 Ragonnet R, Flegg JA, Brilleman SL, Tiemersma EW, Melsew YA, McBryde ES et al. Revisiting the natural history of pulmonary tuberculosis: A bayesian estimation of natural recovery and mortality rates. Clin Infect Dis 2020. doi:10.1093/cid/ciaa602.

17 Blahd M, Leslie EI, Rosenthal SR. Infectiousness of the ‘closed case’ in tuberculosis. Am J Public Health Nations Health 1946; 36: 723–726.

18 National Tuberculosis Institute B. Tuberculosis in a rural population of south india: A five-year epidemiological study. Bull World Health Organ 1974; 51: 473–88.

19 Onozaki I, Law I, Sismanidis C, Zignol M, Glaziou P, Floyd K. National tuberculosis prevalence surveys in asia, 1990-2012: An overview of results and lessons learned. Trop Med Int Health 2015; 20: 1128–1145.

20 Mungai BN, Joekes E, Masini E, Obasi A, Manduku V, Mugi B et al. ‘If not TB, what could it be?’ Chest x-ray findings from the 2016 kenya tuberculosis prevalence survey. Thorax 2021; 76: 607–614.

21 Tiemersma EW, Werf MJ van der, Borgdorff MW, Williams BG, Nagelkerke NJD. Natural history of tuberculosis: Duration and fatality of untreated pulmonary tuberculosis in HIV negative patients: A systematic review. PLoS ONE 2011; 6: e17601.

22 Murray LM. Bayesian state-space modelling on high-performance hardware using LibBi. 13063277 [stat] 2013.http://arxiv.org/abs/1306.3277 (accessed 6 Aug2021).

23 R Core Team. R: A language and environment for statistical computing. R Foundation for Statistical Computing: Vienna, Austria, 2020 https://www.R-project.org/.

24 Wang L, Zhang H, Ruan Y, Chin DP, Xia Y, Cheng S et al. Tuberculosis prevalence in china, 1990-2010; a longitudinal analysis of national survey data. Lancet 2014; 383: 2057–64.

25 Getahun H, Gunneberg C, Granich R, Nunn P. HIV infection-associated tuberculosis: The epidemiology and the response. Clinical infectious diseases : an official publication of the Infectious Diseases Society of America 2010; 50 Suppl 3: S201–7.

26 Jeon CY, Murray MB. Diabetes mellitus increases the risk of active tuberculosis: A systematic review of 13 observational studies. PLoS Medicine 2008; 5: e152.

27 Lonnroth K, Williams BG, Cegielski P, Dye C. A consistent log-linear relationship between tuberculosis incidence and body mass index. International Journal of Epidemiology 2010; 39: 149–155.

28 Holmes CB, Hausler H, Nunn P. A review of sex differences in the epidemiology of tuberculosis. The International Journal of Tuberculosis and Lung Disease 1998; 2: 96–104.

29 Menzies NA, Wolf E, Connors D, Bellerose M, Sbarra AN, Cohen T et al. Progression from latent infection to active disease in dynamic tuberculosis transmission models: A systematic review of the validity of modelling assumptions. Lancet Infect Dis 2018; 18: e228–e238.

30 Storla DG, Yimer S, Bjune GA. A systematic review of delay in the diagnosis and treatment of tuberculosis. BMC public health 2008; 8: 15.

31 Ku C-C, MacPherson P, Khundi M, Nzawa R, Feasey HR, Nliwasa M et al. Estimated durations of asymptomatic, symptomatic, and care-seeking phases of tuberculosis disease. medRxiv 2021; : 2021.03.17.21253823.

32 Glaziou P, Dodd PJ, Dean A, Floyd K. Methods used by WHO to estimate the global burden of TB disease. 2020.

33 Kendall EA, Shrestha S, Dowdy DW. The epidemiological importance of subclinical tuberculosis: A critical re-appraisal. Am J Respir Crit Care Med 2020. doi:10.1164/rccm.202006-2394PP.

