## Supplementary materials for "The natural history of TB disease-a synthesis of data to quantify progression and regression across the spectrum"

### Table of Contents

**S1 Disease model structure** **1**

**S2 Open TB vs Clinical TB** **2**

**S3 Symptomatic Minimal** **4**

**S4 Data inclusion and exclusion** **5**
S4.1 Data types 5
S4.2 Timings 5
S4.3 Included data 6
S4.4 Excluded data 18

**S5 Fitting process** **22**
S5.1 Weighting 24
S5.2 Duration of disease 25
S5.3 Prevalence ratios 26
S5.4 True Minimals 27

**S6 Minimal disease** **29**

**S7 Disease pathways** **30**

**S8 Duration of symptoms** **33**

**S9 Additional results** **36**

**S10 Sensitivity Analyses** **42**
S10.1 Fitting 42
S10.2 Cohort model 46

***References*** ***49***

### S1 Disease model structure

Our model structure focuses on the spectrum of disease, with infection and progression from infection already assumed to have happened. We split the progression into three states, as shown in figure 1; minimal, subclinical, and clinical disease.

Minimal disease is the earliest stage of disease from infection, being non-infectious, but with pathological changes to the lung visible on, originally, chest x-ray but also other forms of chest imaging such as computed tomography (CT). Along with being the first stage of disease after infection, minimal is the final stage before recovery, with regression back to minimal possible within the spectrum framework, and then natural recovery from disease possible.

In the forward progression, the stage after minimal is subclinical. This is an infectious disease state, but without sufficient symptoms to present for screening. In other words, this is an infectious but asymptomatic state. Within the spectrum, there is progression to clinical disease (i.e. development of symptoms) and regression to minimal disease (i.e. becoming non-infectious) from subclinical disease.

Clinical disease, symptomatic and infectious, is the final disease state. This state can only be reached by passing through minimal and subclinical first. As with the other two states, there are two transition possibilities out of clinical disease, regression to subclinical disease (i.e. resolution of symptoms but remaining infectious), and death from TB.

In the model structure, there is possibility to both progress and regress, but in visualising the model, such as figure 1, arrows that point right indicate the disease progressing to a more severe state, and arrows that point left indicate the disease regressing to a less severe state.

| Disease State | Minimal | Subclinical | Clinical |
|---|---|---|---|
| **Clinical description** | Radiological changes attributable to TB but sputum negative (non-infectious) | Sputum positive TB disease that does not pass a symptom screen (infectious) | Symptomatic and sputum positive TB diseaese (infectious) |
| **Systematic review tests** | Chest x-ray or fluoroscopy positive<br>Smear or culture negative | Smear or culture positive<br>Symptom negative | Smear or culture positive<br>Symptom positive |
| **Systematic review notation** | cxr.pos<br>micro.neg<br>symp.pos/neg/unk | cxr.pos<br>micro.pos<br>symp.neg | cxr.pos<br>micro.pos<br>symp.pos |

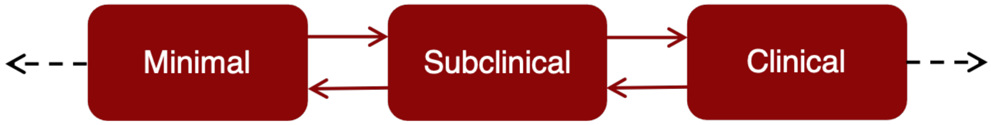


*Figure 1: TB natural history model. States and transitions in red are considered in the model fitting, with dashed parameters holding a fixed value and solid transitions being estimated from the data. Each state is defined by a clinical description and the tests that would have been used in clinical settings at the time our data was recorded with the notation used for data collection in table S4 described in the final row. The black dashed lines out from minimal and clinical are recovery and TB mortality respectively.*

### S2 Open TB vs Clinical TB

In a systematic review, Tiemersma et al describe how study results were interpreted as having people with either smear negative or smear positive disease:[1]

> *in studies where patients were described as having "open" tuberculosis or "bacillary tuberculosis" before 1930 (when culture became available) we assumed that these patients were smear-positive.*

This definition is maintained in the work of Ragonnet et al.[2]

As we were not attempting to stratify bacteriological status any further by smear status, we instead interpreted bacillary tuberculosis as simply bacteriologically positive. The definition of ''open'' tuberculosis was less clear cut, with different studies choosing different explanations.

Szucs, in 1926, defines open and closed tuberculosis based solely on the symptom status of the individual:[3]

> *Among others, we classify tuberculous patients as open and closed cases, based on the presence or absence of expectoration.*

However, they point out that absence of symptoms does not indicate a negative bacteriological status, in essence diagnosing their patients with subclinical disease using more modern terminology.

> *We succeeded in demonstrating the presence of tubercle bacilli in the spray of two of our recently admitted patients in the absence of any expectoration.*

Another study in 1947 provides a different definition for open and closed. Tattersall compares his work with that of Lindhardt in Denmark, stating that:[4,5]

> *These results accord closely with Lindhardt's finding in Denmark during the same period, which enhances the value of comparison of the present series of sputum +ve cases with the results of the Danish survivals of open cases.*

This suggests that rather than open being equivalent to clinical and closed being equivalent to subclinical, open is actually all bacteriologically positive disease (i.e. subclinical and clinical).

Finally, Blahd, in 1946 states that Illinois state defines that all cases that were bacteriologically positive must be defined as open TB, using the same definition as Tattersall.[6] However he also considers the possibility of infection from closed TB.

> *All cases in which a positive sputum has been shown must by Illinois law be considered open for a period of at least three months and thereafter until three successive specimens of sputum, collected at intervals of one week, contain no tubercle bacilli… Although a so-called "closed" case is not as grave a source of infection as a frankly open one, it is more insidious.*

Our definition of clinical aligns closely with the National Tuberculosis Association's definition of "active" TB, as described in their diagnostic standards from 1940.[7] By this point there was less focus on open and closed definitions and more consideration of symptoms and bacteriology together:

> *Symptoms unchanged, worse or less severe, but not completely abated. Lesions not completely healed or progressive according X-ray examination. Sputum almost always contains tubercle bacilli.*

Overall, the definitions of "open" tuberculosis and the final definition of "active" tuberculosis do not differ significantly, and again are not dissimilar to our definition of clinical disease. Therefore, where Ragonnet et al have provided a TB mortality rate for smear positive TB, we are instead using this number as the TB mortality rate from clinical TB disease.[2]

### S3 Symptomatic Minimal

Minimal disease was classified as when an individual had radiological changes attributable to TB but negative bacteriology, regardless of symptoms. Although progressing from a potentially symptomatic bacteriologically negative state to an asymptomatic bacteriologically positive seems unlikely, a number of sources suggested that there was no need to consider an alternative progression for symptomatic minimal.

Firstly, there is insufficient data to show an obvious split in progression between symptomatic and asymptomatic bacteriologically negative individuals (see figure 2), the diagnostic standards from the 1940s placed significantly less weight on symptoms if they were not accompanied by a positive sputum, and the prevalence survey in Cambodia in 2002 found that symptoms in culture negative individuals were not associated with future bacteriological positivity.[7,8] We also know that TB symptoms are highly non-specific, and so there is no guarantee that symptoms occurring whilst an individual has minimal disease are actually caused by TB and not by something else.

Therefore, we have considered all bacteriologically negative individuals to be minimal, regardless of symptom status.

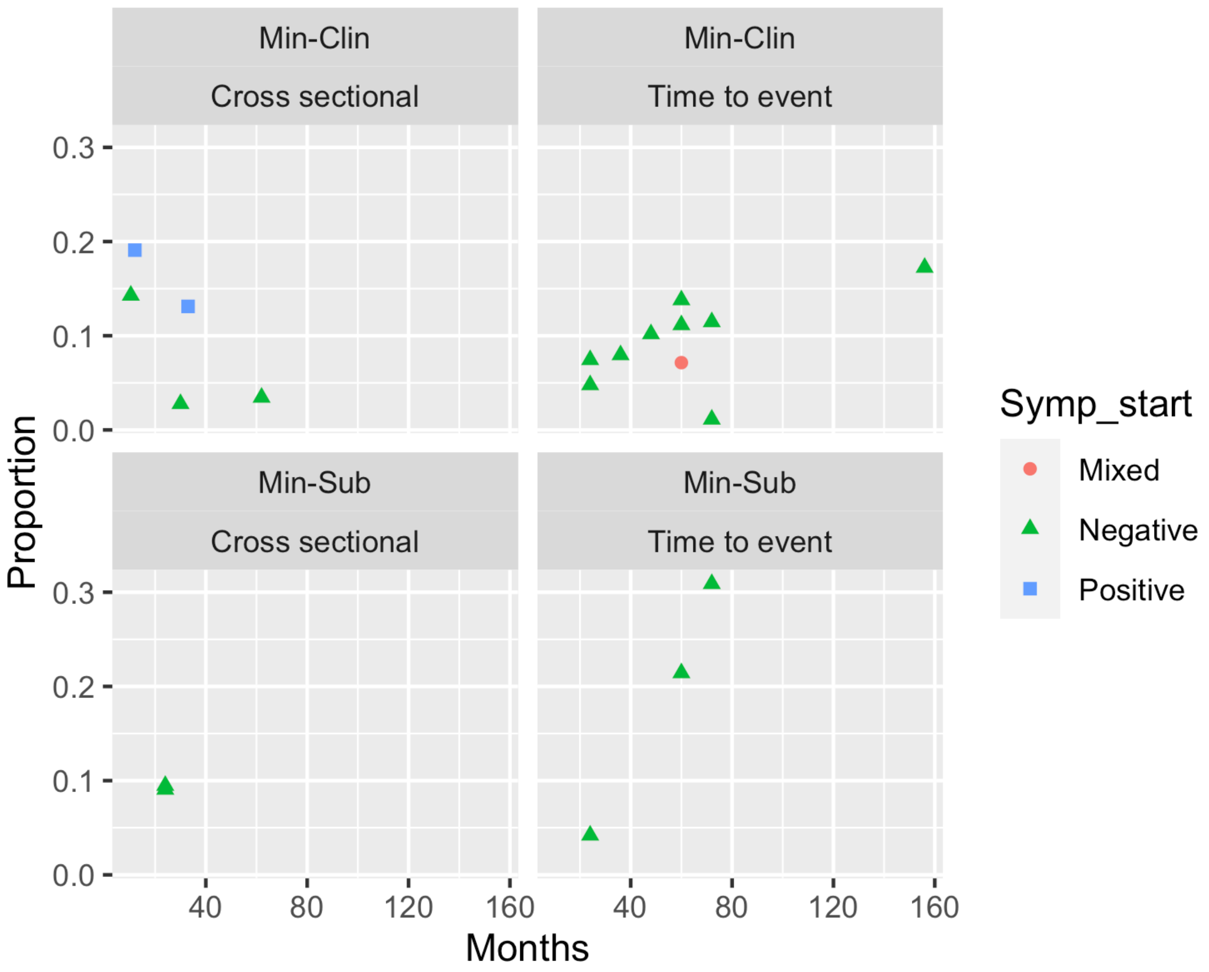


*Figure 2: The different progression rates for minimal disease, based on symptoms, transition, and record type*

### S4 Data inclusion and exclusion

#### S4.1 Data types

As explained in the main text, there were two study types included. In table 1 we report the data types for each line of data, in column "Follow-up method". There were 38 data points reported as cumulative, and 16 reported as cross sectional.

#### S4.2 Timings

For cross-sectional data, if an average follow-up time was given, that is the time used. Otherwise, if a minimum and maximum follow-up time was given, the times have been summed and halved to give the follow-up time used. For time-to-event data, the maximum follow-up time given was used. The times in table 1, column "Months of follow-up" reflect these choices.

#### S4.3 Included data

All data that was included in the final data fit is in table 1Table 1: A table on all the studies included, the model transitions they parameterise, and, where applicable, notes on why a certain decision has been taken. The number of repeats column reports the number of data points that a study provides within a single transition. This is then used to calculate the effective number of people transitioned and the effective cohort size (divides the true numbers which are then rounded to the nearest integer) to appropriately weight each data point so that each study is considered as one.

| First Author | Study ID | Study Continent | Year | Start states | End states | Number transitioned | Cohort size | Months of follow-up | Follow-up method | Model transition | Number of repeats | Effective number transitioned | Effective cohort size | Notes |
|---|---|---|---|---|---|---|---|---|---|---|---|---|---|---|
| Downes | 9 | North America | 1935 | cxr.pos micro.pos sympt.pos | cxr.pos micro.neg sympt.neg | 27 | 342 | 12.0 | cumulative | Clin-Min | 5 | 5 | 68 | |
| Downes | 9 | North America | 1935 | cxr.pos micro.pos sympt.pos | cxr.pos micro.neg sympt.neg | 104 | 342 | 24.0 | cumulative | Clin-Min | 5 | 21 | 68 | |
| Downes | 9 | North America | 1935 | cxr.pos micro.pos sympt.pos | cxr.pos micro.neg sympt.neg | 140 | 342 | 36.0 | cumulative | Clin-Min | 5 | 28 | 68 | |
| Downes | 9 | North America | 1935 | cxr.pos micro.pos sympt.pos | cxr.pos micro.neg sympt.neg | 158 | 342 | 48.0 | cumulative | Clin-Min | 5 | 32 | 68 | |

| First Author | Study ID | Study Continent | Year | Start states | End states | Number transitioned | Cohort size | Months of follow-up | Follow-up method | Model transition | Number of repeats | Effective number transitioned | Effective cohort size | Notes |
|---|---|---|---|---|---|---|---|---|---|---|---|---|---|---|
| Downes | 9 | North America | 1935 | cxr.pos micro.pos sympt.pos | cxr.pos micro.neg sympt.neg | 171 | 342 | 60.0 | cumulative | Clin-Min | 5 | 34 | 68 | |
| Beeuwkes | 10 | North America | 1938 | cxr.pos micro.neg sympt.neg | cxr.unk micro.pos sympt.pos | 16 | f122 | 33.0 | cross sectional | Min-Clin | 1 | 16 | 122 | merged 2 groups exhibiting same start and end |
| Beeuwkes | 10 | North America | 1938 | cxr.pos micro.pos sympt.pos | cxr.unk micro.neg sympt.unk | 10 | 28 | 33.0 | cross sectional | Clin-Min | 1 | 10 | 28 | |
| Puffer | 11 | North America | 1943 | cxr.pos micro.neg sympt.neg | cxr.pos micro.pos sympt.pos | 19 | 528 | 62.0 | cross sectional | Min-Clin | 1 | 19 | 528 | merged 2 groups exhibiting same start and end |
| Puffer | 11 | North America | 1943 | cxr.pos micro.pos sympt.pos | cxr.pos micro.neg sympt.neg | 92 | 384 | 62.0 | cross sectional | Clin-Min | 1 | 92 | 384 | |

| First Author | Study ID | Study Continent | Year | Start states | End states | Number transitioned | Cohort size | Months of follow-up | Follow-up method | Model transition | Number of repeats | Effective number transitioned | Effective cohort size | Notes |
|---|---|---|---|---|---|---|---|---|---|---|---|---|---|---|
| Orrego Puelma | 12 | South America | 1945 | cxr.pos micro.neg sympt.unk | cxr.pos micro.pos sympt.unk | 18 | 67 | 24.0 | cross sectional | Min-Inf | 1 | 18 | 67 | |
| Bobrowitz | 13,14 | North America | 1945 | cxr.pos micro.neg sympt.unk | cxr.pos micro.pos sympt.unk | 26 | 191 | 60.0 | cross sectional | Min-Inf | 1 | 26 | 191 | |
| Lincoln | 16,17 | North America | 1947 | cxr.pos micro.mixed sympt.unk | cxr.pos micro.neg sympt.unk | 45 | 134 | 24.0 | cumulative | Clin-Min | 6 | 8 | 22 | majority tested were micro pos and all reported as active baseed on NTA definitions |
| Lincoln | 16,17 | North America | 1947 | cxr.pos micro.mixed sympt.unk | cxr.pos micro.neg sympt.unk | 71 | 134 | 36.0 | cumulative | Clin-Min | 6 | 12 | 22 | majority tested were micro pos and all reported as |

| First Author | Study ID | Study Continent | Year | Start states | End states | Number transitioned | Cohort size | Months of follow-up | Follow-up method | Model transition | Number of repeats | Effective number transitioned | Effective cohort size | Notes |
|---|---|---|---|---|---|---|---|---|---|---|---|---|---|---|
| | | | | | | | | | | | | | | active baseed on NTA definitions |
| Lincoln | 16,17 | North America | 1947 | cxr.pos micro.mixed sympt.unk | cxr.pos micro.neg sympt.unk | 80 | 134 | 48.0 | cumulative | Clin-Min | 6 | 13 | 22 | majority tested were micro pos and all reported as active baseed on NTA definitions |
| Lincoln | 16,17 | North America | 1947 | cxr.pos micro.mixed sympt.unk | cxr.pos micro.neg sympt.unk | 83 | 134 | 60.0 | cumulative | Clin-Min | 6 | 14 | 22 | majority tested were micro pos and all reported as active baseed on NTA definitions |

| First Author | Study ID | Study Continent | Year | Start states | End states | Number transitioned | Cohort size | Months of follow-up | Follow-up method | Model transition | Number of repeats | Effective number transitioned | Effective cohort size | Notes |
|---|---|---|---|---|---|---|---|---|---|---|---|---|---|---|
| Lincoln | 16,17 | North America | 1947 | cxr.pos micro.mixed sympt.unk | cxr.pos micro.neg sympt.unk | 86 | 134 | 72.0 | cumulative | Clin-Min | 6 | 14 | 22 | majority tested were micro pos and all reported as active baseed on NTA definitions |
| Lincoln | 16,17 | North America | 1947 | cxr.pos micro.mixed sympt.unk | cxr.pos micro.neg sympt.unk | 87 | 134 | 84.0 | cumulative | Clin-Min | 6 | 15 | 22 | majority tested were micro pos and all reported as active baseed on NTA definitions |
| Lincoln | 16,17 | North America | 1947 | cxr.pos micro.neg sympt.unk | cxr.pos micro.relapse sympt.unk | 15 | 314 | 24.0 | cumulative | Min-Clin | 5 | 3 | 63 | majority tested were micro pos and all |

| First Author | Study ID | Study Continent | Year | Start states | End states | Number transitioned | Cohort size | Months of follow-up | Follow-up method | Model transition | Number of repeats | Effective number transitioned | Effective cohort size | Notes |
|---|---|---|---|---|---|---|---|---|---|---|---|---|---|---|
| | | | | | | | | | | | | | | reported as active baseed on NTA definitions |
| Lincoln | 16,17 | North America | 1947 | cxr.pos micro.neg sympt.unk | cxr.pos micro.relapse sympt.unk | 25 | 314 | 36.0 | cumulative | Min-Clin | 5 | 5 | 63 | |
| Lincoln | 16,17 | North America | 1947 | cxr.pos micro.neg sympt.unk | cxr.pos micro.relapse sympt.unk | 32 | 314 | 48.0 | cumulative | Min-Clin | 5 | 6 | 63 | |
| Lincoln | 16,17 | North America | 1947 | cxr.pos micro.neg sympt.unk | cxr.pos micro.relapse sympt.unk | 35 | 314 | 60.0 | cumulative | Min-Clin | 5 | 7 | 63 | |
| Lincoln | 16,17 | North America | 1947 | cxr.pos micro.neg sympt.unk | cxr.pos micro.relapse sympt.unk | 36 | 314 | 72.0 | cumulative | Min-Clin | 5 | 7 | 63 | |
| Alling | 16,19 | North America | 1948 | cxr.pos micro.neg | cxr.pos micro.relapse | 8 | 58 | 60.0 | cumulative | Min-Clin | 2 | 4 | 29 | |

| First Author | Study ID | Study Continent | Year | Start states | End states | Number transitioned | Cohort size | Months of follow-up | Follow-up method | Model transition | Number of repeats | Effective number transitioned | Effective cohort size | Notes |
|---|---|---|---|---|---|---|---|---|---|---|---|---|---|---|
| | | | | sympt.unk | sympt.unk | | | | | | | | | |
| Alling | 16,19 | North America | 1948 | cxr.pos<br>micro.neg<br>sympt.unk | cxr.pos<br>micro.relapse<br>sympt.unk | 10 | 58 | 156.0 | cumulative | Min-Clin | 2 | 5 | 29 | |
| Marshall | 18 | Europe | 1948 | cxr.pos<br>micro.pos<br>sympt.pos | cxr.pos<br>micro.neg<br>sympt.unk | 2 | 52 | 6.0 | cumulative | Clin-Min | 1 | 2 | 52 | |
| Borgen | 20,21 | Europe | 1949 | cxr.pos<br>micro.neg<br>sympt.pos | cxr.pos<br>micro.pos<br>sympt.pos | 4 | 144 | 30.0 | cross sectional | Min-Clin | 1 | 4 | 144 | merged 2 groups exhibiting same start and end |
| Manser | 22 | Europe | 1951 | cxr.pos<br>micro.pos<br>sympt.unk | cxr.pos<br>micro.neg<br>sympt.unk | 15 | 40 | 6.0 | cross sectional | Clin-Min | 1 | 15 | 40 | |
| Breu | 23 | Europe | 1952 | cxr.pos<br>micro.neg<br>sympt.unk | cxr.pos<br>micro.pos<br>sympt.unk | 48 | 904 | 25.5 | cross sectional | Min-Inf | 1 | 48 | 904 | |

| First Author | Study ID | Study Continent | Year | Start states | End states | Number transitioned | Cohort size | Months of follow-up | Follow-up method | Model transition | Number of repeats | Effective number transitioned | Effective cohort size | Notes |
|---|---|---|---|---|---|---|---|---|---|---|---|---|---|---|
| Sikand | 24 | Asia | 1958 | cxr.pos micro.neg sympt.unk | cxr.pos micro.pos sympt.unk | 38 | 319 | 12.0 | cumulative | Min-Inf | 1 | 38 | 319 | |
| Tuberculosis Society of Scotland | #6897 | Europe | 1959 | cxr.pos micro.neg sympt.neg | cxr.pos micro.pos sympt.unk | 9 | 95 | 24.0 | cross sectional | Min-Sub | 1 | 9 | 95 | assume lack of sysmptom persists |
| Frimodt-Moller | 27 | Asia | 1961 | cxr.pos micro.neg sympt.unk | cxr.pos micro.pos sympt.unk | 11 | 86 | 12.0 | cumulative | Min-Inf | 3 | 4 | 29 | |
| Frimodt-Moller | 27 | Asia | 1961 | cxr.pos micro.neg sympt.unk | cxr.pos micro.pos sympt.unk | 18 | 86 | 24.0 | cumulative | Min-Inf | 3 | 6 | 29 | |
| Frimodt-Moller | 27 | Asia | 1961 | cxr.pos micro.neg sympt.unk | cxr.pos micro.pos sympt.unk | 25 | 86 | 36.0 | cumulative | Min-Inf | 3 | 8 | 29 | |
| Pamra | 37 | Asia | 1968 | cxr.pos micro.neg sympt.neg | cxr.pos micro.pos sympt.pos | 2 | 178 | 72.0 | cumulative | Min-Clin | 1 | 2 | 178 | |

| First Author | Study ID | Study Continent | Year | Start states | End states | Number transitioned | Cohort size | Months of follow-up | Follow-up method | Model transition | Number of repeats | Effective number transitioned | Effective cohort size | Notes |
|---|---|---|---|---|---|---|---|---|---|---|---|---|---|---|
| Pamra | 37 | Asia | 1968 | cxr.pos micro.neg sympt.neg | cxr.pos micro.pos sympt.neg | 55 | 178 | 72.0 | cumulative | Min-Sub | 1 | 55 | 178 | |
| National Tuberculosis Insitute | 28-36 | Asia | 1968 | cxr.pos micro.neg sympt.unk | cxr.pos micro.pos sympt.unk | 23 | 329 | 18.0 | cross sectional | Min-Inf | 2 | 12 | 165 | |
| National Tuberculosis Insitute | 28-36 | Asia | 1968 | cxr.pos micro.neg sympt.unk | cxr.pos micro.pos sympt.unk | 36 | 271 | 60.0 | cross sectional | Min-Inf | 2 | 18 | 136 | |
| National Tuberculosis Insitute | 28-36 | Asia | 1968 | cxr.pos micro.pos sympt.unk | cxr.pos micro.neg sympt.unk | 86 | 269 | 18.0 | cross sectional | Inf-Min | 2 | 43 | 135 | |
| National Tuberculosis Insitute | 28-36 | Asia | 1968 | cxr.pos micro.pos sympt.unk | cxr.pos micro.neg sympt.unk | 70 | 178 | 36.0 | cross sectional | Inf-Min | 2 | 35 | 89 | |
| Aneja | 38 | Asia | 1977 | cxr.pos micro.neg sympt.pos | cxr.pos micro.pos sympt.unk | 21 | 110 | 12.0 | cross sectional | Min-Clin | 1 | 21 | 110 | assume symptoms persist |

| First Author | Study ID | Study Continent | Year | Start states | End states | Number transitioned | Cohort size | Months of follow-up | Follow-up method | Model transition | Number of repeats | Effective number transitioned | Effective cohort size | Notes |
|---|---|---|---|---|---|---|---|---|---|---|---|---|---|---|
| Hong Kong Chest Service | 39-42 | Asia | 1981 | cxr.pos micro.neg sympt.mixed | cxr.pos micro.pos sympt.unk | 40 | 176 | 3.0 | cumulative | Min-Inf | 8 | 5 | 22 | |
| Hong Kong Chest Service | 39-42 | Asia | 1981 | cxr.pos micro.neg sympt.mixed | cxr.pos micro.pos sympt.unk | 49 | 176 | 6.0 | cumulative | Min-Inf | 8 | 6 | 22 | |
| Hong Kong Chest Service | 39-42 | Asia | 1981 | cxr.pos micro.neg sympt.mixed | cxr.pos micro.pos sympt.unk | 61 | 176 | 12.0 | cumulative | Min-Inf | 8 | 8 | 22 | |
| Hong Kong Chest Service | 39-42 | Asia | 1981 | cxr.pos micro.neg sympt.mixed | cxr.pos micro.pos sympt.unk | 67 | 176 | 18.0 | cumulative | Min-Inf | 8 | 8 | 22 | |
| Hong Kong Chest Service | 39-42 | Asia | 1981 | cxr.pos micro.neg sympt.mixed | cxr.pos micro.pos sympt.unk | 69 | 176 | 24.0 | cumulative | Min-Inf | 8 | 9 | 22 | |
| Hong Kong Chest Service | 39-42 | Asia | 1981 | cxr.pos micro.neg sympt.mixed | cxr.pos micro.pos sympt.unk | 70 | 176 | 30.0 | cumulative | Min-Inf | 8 | 9 | 22 | |

| First Author | Study ID | Study Continent | Year | Start states | End states | Number transitioned | Cohort size | Months of follow-up | Follow-up method | Model transition | Number of repeats | Effective number transitioned | Effective cohort size | Notes |
|---|---|---|---|---|---|---|---|---|---|---|---|---|---|---|
| Hong Kong Chest Service | 39-42 | Asia | 1981 | cxr.pos micro.neg sympt.mixed | cxr.pos micro.pos sympt.unk | 71 | 176 | 36.0 | cumulative | Min-Inf | 8 | 9 | 22 | |
| Hong Kong Chest Service | 39-42 | Asia | 1981 | cxr.pos micro.neg sympt.mixed | cxr.pos micro.pos sympt.unk | 71 | 176 | 60.0 | cumulative | Min-Inf | 8 | 9 | 22 | |
| Cowie | 43 | Africa | 1984 | cxr.pos micro.neg sympt.unk | cxr.pos micro.pos sympt.unk | 88 | 152 | 58.0 | cumulative | Min-Inf | 1 | 88 | 152 | |
| Norregaard | 45 | Europe | 1985 | cxr.pos micro.neg sympt.mixed | cxr.pos micro.pos sympt.neg | 6 | 28 | 60.0 | cumulative | Min-Sub | 1 | 6 | 28 | |
| Norregaard | 45 | Europe | 1985 | cxr.pos micro.neg sympt.mixed | cxr.pos micro.pos sympt.pos | 2 | 28 | 60.0 | cumulative | Min-Clin | 1 | 2 | 28 | |
| Anastasatu | 44 | Europe | 1985 | cxr.pos micro.neg sympt.neg | cxr.pos micro.pos sympt.unk | 6 | 143 | 24.0 | cumulative | Min-Sub | 1 | 6 | 143 | assume lack of sysmptom persists |

| First Author | Study ID | Study Continent | Year | Start states | End states | Number transitioned | Cohort size | Months of follow-up | Follow-up method | Model transition | Number of repeats | Effective number transitioned | Effective cohort size | Notes |
|---|---|---|---|---|---|---|---|---|---|---|---|---|---|---|
| Okada | 8 | Asia | 2004 | cxr.pos micro.neg sympt.neg | cxr.pos micro.pos sympt.unk | 28 | 309 | 24.0 | cross sectional | Min-Sub | 1 | 28 | 309 | split group by symptoms dependent on proportion found in prevalence survey |
| Okada | 8 | Asia | 2004 | cxr.pos micro.neg sympt.neg | cxr.pos micro.pos sympt.pos | 23 | 309 | 24.0 | cumulative | Min-Clin | 1 | 23 | 309 | added in 39% (18) of those picked up at 2 year follow-up using known subclinical proportion at prevalence surveys |

### S4.4 Excluded data

As can be seen in table 2, some of the data that was originally extracted for the wider review was not eligible for this work. In total 20 cohorts were excluded, from from 10 different studies. Most (22) rows of data that were excluded, had an initial state with x-ray negative, and one study was observing a cohort who, although already x-ray positive, were not expected to progress to infectious TB disease. Others were excluded for too much uncertainty within the start and end states, or changes only within states, such as change in x-ray severity, but no change in bacteriological or symptom status. These reasonings are laid out in table 2

*Table 2: A table on all the studies excluded,and reasons why they have not been included*

| author | record.id | cohort.end | continent | box.start | box.end | n.end | cohort.size | x.months | notes |
|---|---|---|---|---|---|---|---|---|---|
| Beeuwkes | 10 | 1938 | North America | cxr.neg micro.neg sympt.neg | cxr.unk micro.pos sympt.pos | 1 | 784 | 33 | initial state x-ray negative |
| Beeuwkes | 10 | 1938 | North America | cxr.neg micro.neg sympt.neg | cxr.unk micro.neg sympt.pos | 3 | 784 | 33 | initial state x-ray negative |
| Beeuwkes | 10 | 1938 | North America | cxr.pos micro.neg sympt.neg | cxr.unk micro.neg sympt.pos | 5 | 79 | 33 | change within state |
| National Tuberculosis Insitute | 28-36 | 1968 | Asia | cxr.neg micro.neg sympt.unk | cxr.pos micro.pos sympt.unk | 44 | 31490 | 18 | initial state x-ray negative |
| National Tuberculosis Insitute | 28-36 | 1968 | Asia | cxr.neg micro.neg sympt.unk | cxr.pos micro.pos sympt.unk | 99 | 17936 | 60 | initial state x-ray negative |
| Borgen | 20,21 | 1949 | Europe | cxr.neg micro.unk sympt.unk | cxr.pos micro.pos sympt.pos | 4 | 6684 | 30 | initial state x-ray negative |
| Madsen | 46 | 1940 | Europe | cxr.neg micro.unk sympt.neg | cxr.pos micro.mixed sympt.unk | 4 | 2071 | 12 | initial state x-ray negative |
| Madsen | 46 | 1940 | Europe | cxr.neg micro.unk | cxr.pos micro.mixed | 7 | 2071 | 24 | initial state x-ray negative |

| author | record.id | cohort.end | continent | box.start | box.end | n.end | cohort.size | x.months | notes |
|---|---|---|---|---|---|---|---|---|---|
| | | | | sympt.neg | sympt.unk | | | | |
| Madsen | 46 | 1940 | Europe | cxr.neg micro.unk sympt.neg | cxr.pos micro.mixed sympt.unk | 10 | 2071 | 36 | initial state x-ray negative |
| Madsen | 46 | 1940 | Europe | cxr.neg micro.unk sympt.neg | cxr.pos micro.mixed sympt.unk | 11 | 2071 | 48 | initial state x-ray negative |
| Madsen | 46 | 1940 | Europe | cxr.neg micro.unk sympt.neg | cxr.pos micro.mixed sympt.unk | 16 | 2071 | 60 | initial state x-ray negative |
| Madsen | 46 | 1940 | Europe | cxr.neg micro.unk sympt.neg | cxr.pos micro.mixed sympt.unk | 17 | 2071 | 72 | initial state x-ray negative |
| Okada | 8 | 2004 | Asia | cxr.neg micro.neg sympt.neg | cxr.pos micro.pos sympt.unk | 32 | 21580 | 24 | initial state x-ray negative |
| Okada | 8 | 2004 | Asia | cxr.pos micro.neg sympt.neg | cxr.neg micro.neg sympt.unk | 26 | 309 | 24 | ends outside disease |
| International Union Against Tuberculosis Committee on Prophylaxis | 47 | 1982 | Europe | cxr.pos micro.neg sympt.unk | cxr.pos micro.pos sympt.unk | 97 | 6990 | 60 | cohort not expected to progress - effectively initial state x-ray negative |
| Sikand | 24 | 1958 | Asia | cxr.neg micro.unk sympt.unk | cxr.pos micro.pos sympt.unk | 89 | 11268 | 43 | initial state x-ray negative |
| Sikand | 24 | 1958 | Asia | cxr.neg micro.unk sympt.unk | cxr.pos micro.neg sympt.unk | 251 | 11268 | 43 | initial state x-ray negative |

| author | record.id | cohort.end | continent | box.start | box.end | n.end | cohort.size | x.months | notes |
|---|---|---|---|---|---|---|---|---|---|
| Groth-Petersen | 48,49 | 1952 | Europe | cxr.neg micro.unk sympt.unk | cxr.pos micro.mixed sympt.unk | 59 | 45953 | 24 | initial state x-ray negative |
| Groth-Petersen | 48,49 | 1952 | Europe | cxr.neg micro.unk sympt.unk | cxr.pos micro.mixed sympt.unk | 108 | 45953 | 48 | initial state x-ray negative |
| Groth-Petersen | 48,49 | 1952 | Europe | cxr.neg micro.unk sympt.unk | cxr.pos micro.mixed sympt.unk | 64 | 116639 | 24 | initial state x-ray negative |
| Groth-Petersen | 48,49 | 1952 | Europe | cxr.neg micro.unk sympt.unk | cxr.pos micro.mixed sympt.unk | 147 | 116639 | 48 | initial state x-ray negative |
| Groth-Petersen | 48,49 | 1952 | Europe | cxr.neg micro.unk sympt.unk | cxr.pos micro.mixed sympt.unk | 73 | 103373 | 48 | initial state x-ray negative |
| Groth-Petersen | 48,49 | 1952 | Europe | cxr.neg micro.unk sympt.unk | cxr.pos micro.mixed sympt.unk | 35 | 69607 | 48 | initial state x-ray negative |
| Groth-Petersen | 48,49 | 1952 | Europe | cxr.pos micro.unk sympt.unk | cxr.pos micro.mixed sympt.unk | 46 | 2877 | 48 | too much uncertainty |
| Groth-Petersen | 48,49 | 1952 | Europe | cxr.pos micro.unk sympt.unk | cxr.pos micro.mixed sympt.unk | 64 | 9693 | 48 | too much uncertainty |
| Groth-Petersen | 48,49 | 1952 | Europe | cxr.pos micro.unk sympt.unk | cxr.pos micro.mixed sympt.unk | 43 | 11366 | 48 | too much uncertainty |
| Groth-Petersen | 48,49 | 1952 | Europe | cxr.pos micro.unk sympt.unk | cxr.pos micro.mixed sympt.unk | 45 | 12400 | 48 | too much uncertainty |

| author | record.id | cohort.end | continent | box.start | box.end | n.end | cohort.size | x.months | notes |
|---|---|---|---|---|---|---|---|---|---|
| Styblo | 50 | 1965 | Europe | cxr.neg<br>micro.neg<br>sympt.neg | cxr.unk<br>micro.pos<br>sympt.unk | 241 | 73000 | 30 | initial state x-ray negative |
| Rubinshteyn | 51 | 1940 | Russia | | | NA | NA | NA | initial state x-ray negative |

### S5 Fitting process

The equations to define the model system are:

$$\frac{dM}{dt} = -m\_s * M + s\_m * S - r * M$$
$$\frac{dS}{dt} = m\_s * M - s\_m * S - s\_c * S + c\_s * C$$
$$\frac{dC}{dt} = s\_c * S - c\_s * C - d * C$$

where:

- $M$, $S$, and $C$ are the states for minimal, subclinical, and clinical respectively
- $m\_s$, $s\_m$, $s\_c$, and $c\_s$ are transitions between the states, where the first letter is the start state and the second letter is the end state
- $r$ is recovery from minimal disease
- $d$ is death from clinical disease (there is no other death included in the model)

As the data described how a cohort changed over time, and described only one outcome, the fitting process used a model system for each of the transitions and and data types, totalling 16 different versions of the model system. Full code is available on GitHub.

We used uniform priors for the four estimated parameters, all with a range of 0 to 12, where 12 would be equivalent to changing state once a month. During the fitting process, potential parameters are trialled within this range. figure 3 shows the different parameter values that were accepted over the 10,000 iterations of the model fit.

These accepted parameters in turn, inform figure 4, which shows the correlation between two parameters. It also shows the overall distribution of the accepted parameters. We see a strong positive correlation between the parameters that control transition between minimal and subclinical; as one transition increases, the also has to increase to prevent there being excess people in one state and too few in another. We also see this with the parameters that control transition between subclinical and clinical.

The rest of the pairings have slightly weaker, negative correlations. This is clearest with the subclinical to minimal and subclinical to clinical pairing, as if one increases, the other has to decrease to make sure that there are still sufficient individuals in subclinical to fit the data.

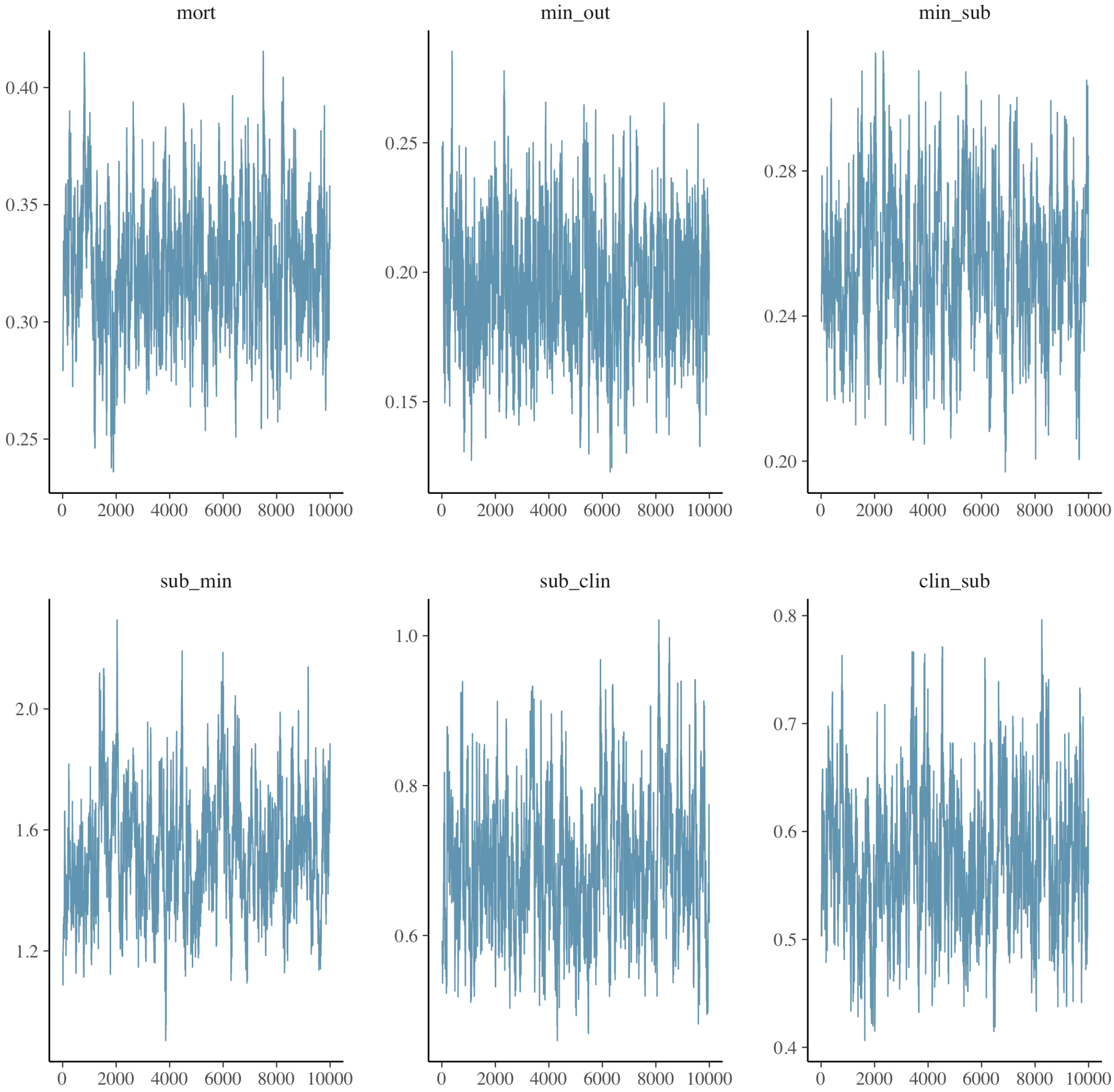


*Figure 3: The traces of the accepted parameter range from the fitting process*

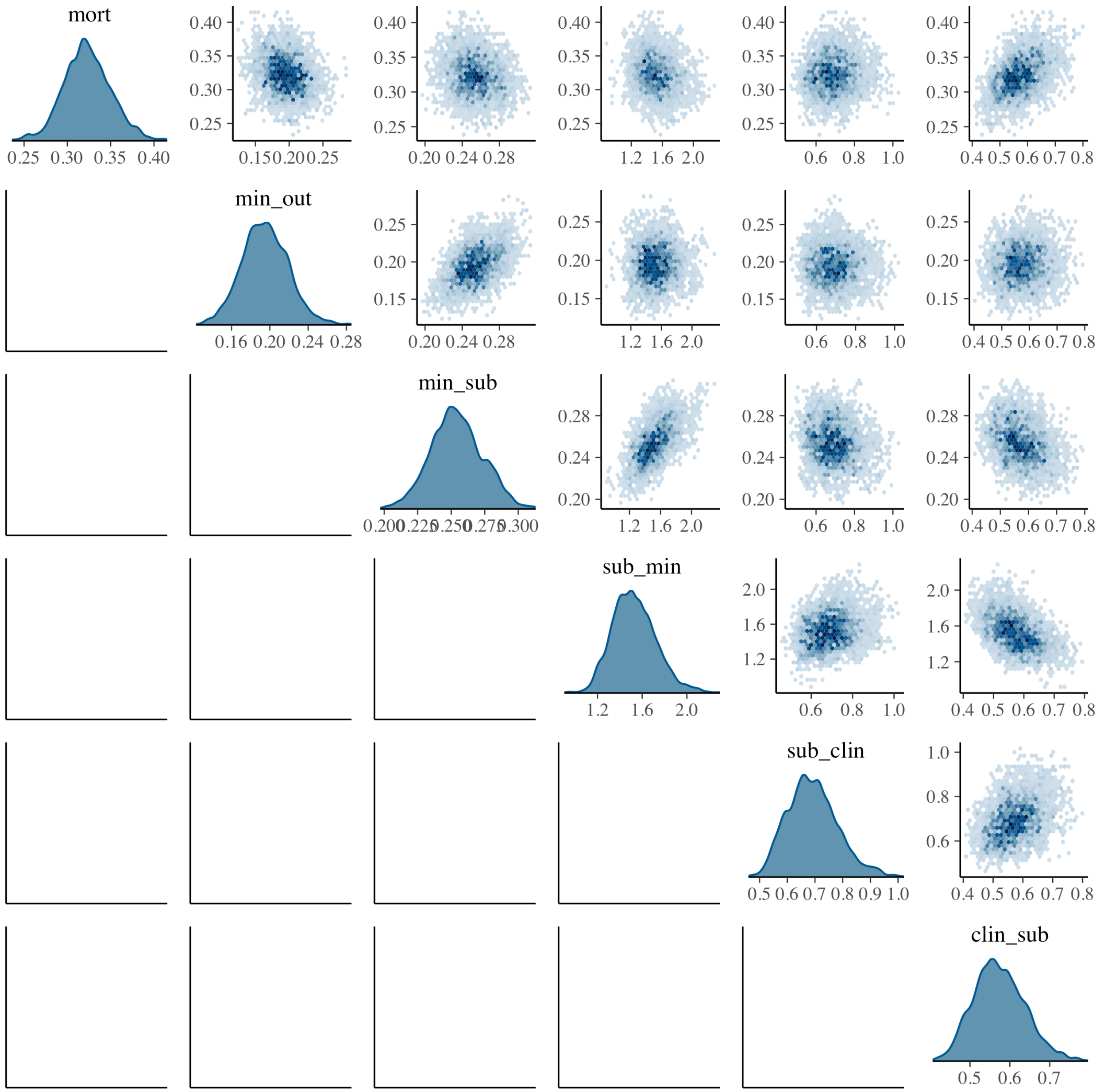


*Figure 4: The distributions of and correlations between the accepted parameters from the fitting process*

### S5.1 Weighting

When calculating the likelihood, larger studies were weighted by the original cohort size to reflect the increased confidence that such studies provide. Thus larger cohorts have a heavier weighting and constrain the model more.

In order to prevent a single study with multiple observations being over-represented in the fit of a transition, we down-weighted both the sample size and the number of people transitioning by the number of repeated observations. This maintains the observed proportion to transition whilst reducing confidence and thus importance given to each individual data point within the study. This is shown in table 1, with the number of repeats and the re-calculated effective cohort size and number transitioning. This ensures that the proportion remains the same, but less weight is given to each individual data point.

#### S5.2 Duration of disease

Tiemersma et al use an assumption of exponential duration of disease to quote an "average" duration of disease as three years and calculates this from the incident cases occuring between each survey.[1,28] They state that a $\delta$ of 0.3 fits the cumulative distribution for the number of observed cases and thus $\frac{1}{\delta} = 3.33$ years is the average duration given by the data, but that missed cases mean that is likely an over-estimate and so 3 years is the average duration of disease . What they are then quoting as average is the mean, so the median duration can be given by $\frac{ln(2)}{\delta}$. Taking $\delta = 0.33$ so that mean duration is 3, gives a median duration of 2.1 years.

The numbers quoted are incident cases between each survey, so we can use the duration of disease looking at a cohort that starts in subclinical disease. Therefore we need to find that at 2 years, 50% of the cohort that started in subclinical, is either still subclinical or is clinical.

To use this as a fitting point, we want to look at time 2 years, and see how close to 50% the number of people in subclinical + clinical from the subclinical cohort is.

An exponential function can be written as $p = a(b^t)$ where $p$ is prevalence and $t$ is time. We know that at $t = 0, p = 1$ so $a = 1$ and the equation simplifies to $p = b^t$.

We want to look at 2 years, find the prevalence, and then from that, calculate the time at which the prevalence would be 0.5. So to calculate b, we set $p = 0.5$ and $t = t_{med}$.

$$
\begin{aligned}
0.5 &= b^{t_{med}} \\
b &= \left(\frac{1}{2}\right)^{\frac{1}{t_{med}}}
\end{aligned}
$$

So the full equation, at the fitting point of $t = 2$ and rearranging for $t_{med}$ gives:

$$
\begin{aligned}
p &= \frac{1^{\frac{2}{t_{med}}}}{2} \\
ln(p) &= ln\left(\frac{1}{2}\right)\frac{2}{t_{med}} \\
t_{med} &= ln\left(\frac{1}{2}\right)\frac{2}{ln(p)} \\
t_{med} &= \big(ln(1) - ln(2)\big)\frac{2}{ln(p)} \\
t_{med} &= -2\frac{ln(2)}{ln(p)}
\end{aligned}
$$

This means, from fitting at a single time point, we can estimate the median duration of disease using the assumption of an exponential distribution of duration.

#### S5.3 Prevalence ratios

Prevalence surveys have found that approximately 50% of people with bacteriologically positive disease do not report experiencing symptoms,.[52] Whilst harder to ascertain, estimates of the proportion of people with bacteriologically negative disease range from two to three times the number of people with infectious disease. Both these have been included in the model fit as data points, calculated from the steady states of the system equations.

The subclinical to clinical ratio is calculated:

$$\begin{aligned} \frac{dC}{dt} &= s\_c * S - c\_s * C - d * C \\ 0 &= s\_c * S - c\_s * C - d * C \\ s\_c * S &= c\_s * C + d * C \\ \frac{S}{C} &= \frac{c\_s + d}{s\_c} \end{aligned}$$

For simplicity, the parameters representing transitions have been simplified to single letters:

- $m\_s$ -> $e$
- $s\_m$ -> $f$
- $s\_c$ -> $g$
- $c\_s$ -> $h$
- $r$ -> $j$
- $d$ -> $k$

and to create a non-zero steady state, an unknown $\alpha$ is the flow of new disease.

The system of equations then becomes:

$$\begin{aligned} \dot{M} &= \alpha - (e + j)M + fS \\ \dot{S} &= eM - (g + f)S + hC \\ \dot{C} &= gS - (h + k)C \end{aligned}$$

Assuming a steady state and using the equation for $\dot{S}$ we can get an equation for M in terms of S and C:

$$\begin{aligned} 0 &= eM - (g + f)S + hC \\ eM &= (g + f)S - hC \\ M &= \frac{(g + f)S - hC}{e} \end{aligned}$$

Substituting C in terms of S:

$$M = \frac{(g+f)S - h\frac{g}{h+k}S}{e}$$

$$M = \frac{(g+f)(h+k) - hg}{e(h+k)}S$$

Then to calculate $\frac{M}{S+C}$:

$$\frac{M}{S+C} = \frac{\frac{(g+f)(h+k) - hg}{e(h+k)}S}{S+C}$$

$$\frac{M}{S+C} = \frac{\frac{(g+f)(h+k) - hg}{e(h+k)}S}{S + \frac{g}{h+k}S}$$

$$\frac{M}{S+C} = \frac{\frac{(g+f)(h+k) - hg}{e(h+k)}}{1 + \frac{g}{h+k}}$$

$$\frac{M}{S+C} = \frac{\frac{(g+f)(h+k) - hg}{e(h+k)}}{\frac{h+k+g}{h+k}}$$

$$\frac{M}{S+C} = \frac{(g+f)(h+k) - hg}{e(h+k+g)}$$

#### S5.4 True Minimals

In the systematic review preceding this work, x-ray positive, bacteriologically negative diseasee was analysed based on reporting of the presumed activity (whether the x-rays were classified as active or inactive). For modelling purposes, there was insufficient data to split groups starting in minimal disease beyond the symptoms at the end and the follow-up collection type, so the distinction between active and inactive x-rays has not been included. However, determining who truly has TB when the only test is an x-ray is difficult. To take this into account, we have used tuberculin skin test (TST) as a proxy for determining whether a positive x-ray is a result of TB infection progressing to disease, and so we can estimate the proportion of people classed as minimal that are actually minimal. These papers were not selected systematically but span a range of time and location. Table 3 shows each of the studies, the number of people who were found to be x-ray positive in the study, and then the number of those who were also TST negative.

*Table 3: The different studies that contributed towards the decision to reduce the proportion of people with positive x-rays that were considered to be truly minimal*

| Author | xray_pos | tst_neg |
|---|---|---|
| Groth-Petersen, 1959 | 37494 | 6097 |
| Roelsgaard, 1964 | 2857 | 772 |

| Author | xray_pos | tst_neg |
|---|---|---|
| Roelsgaard, 1961 | 559 | 169 |
| Scheel, 1937 | 255 | 54 |
| National Tuberculosis Institute, 1974 | 3761 | 1848 |

Applying a meta analysis to this, we find that the fixed effects result is 20% of x-ray positives are TST negative and so unlikely to be caused by TB, and the random effects suggests 28%, as can be seen in figure 5. Therefore throughout this paper we have assumed that 25% of all x-ray positives are non-TB, and thus reduced every cohort that starts in minimal accordingly.

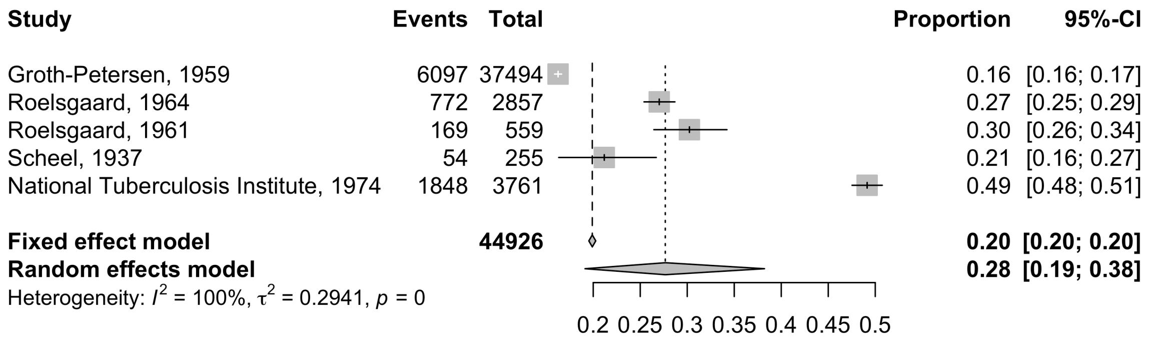


*Figure 5: The results of a meta-analysis on the proportion of those with positive x-rays who also test tst negative, as a proxy for the proportion of positive x-rays that are not caused by TB*

To check this assumption, we have tested this with 0% and 40% of x-ray positives being non-TB, as can be seen in table 6.

### S6 Minimal disease

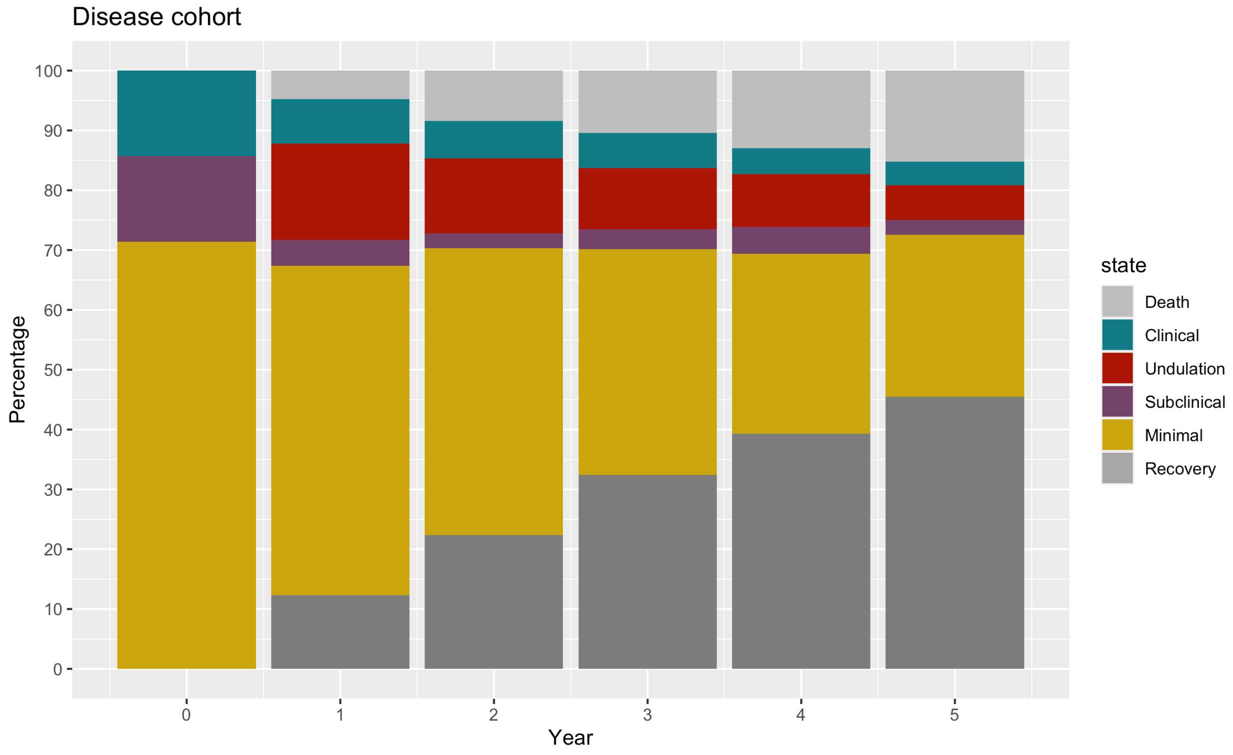


*Figure 6: Trajectories of disease over time given different cohort starts*

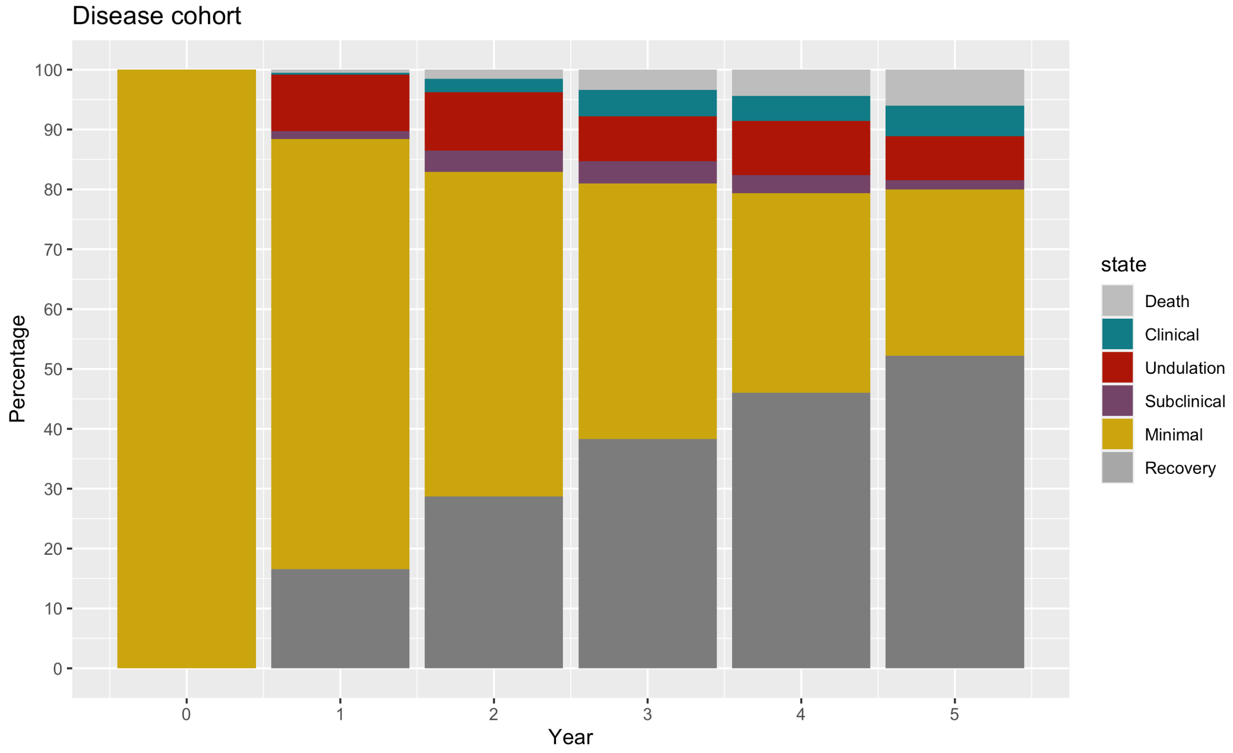


*Figure 7: Trajectories of disease over time given different cohort starts*

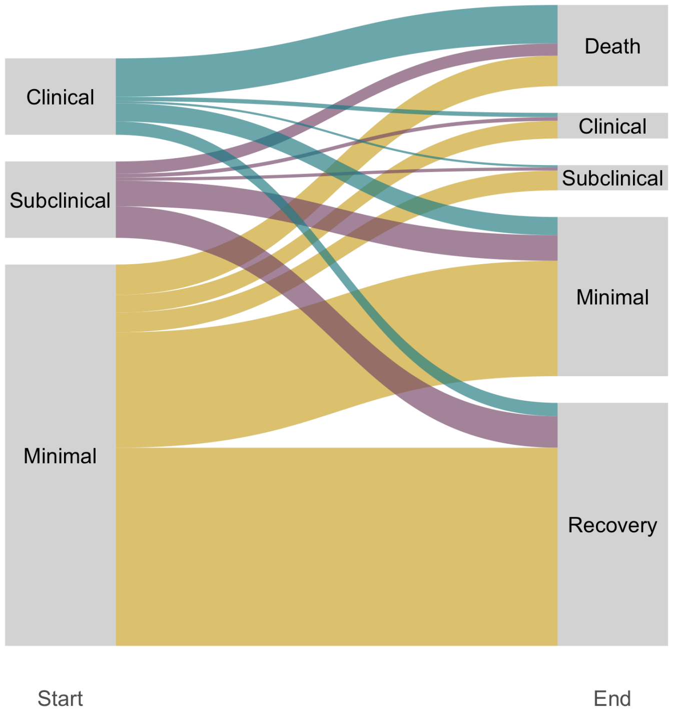


*Figure 8: Final state after five years of people starting in subclinical and clinical disease*

### S7 Disease pathways

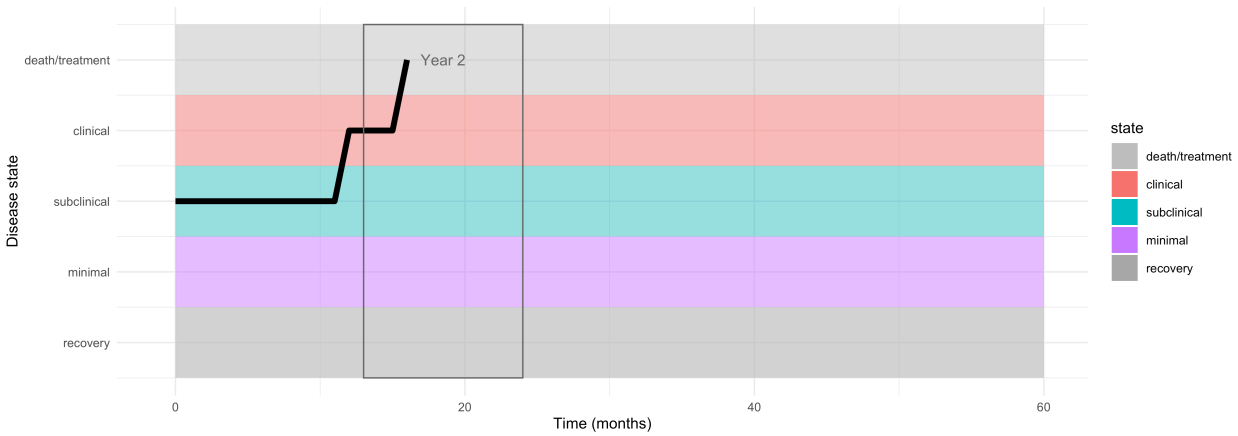


*Figure 9: An example trajectory where the individual dies from TB. In this case, the death happens in year 2*

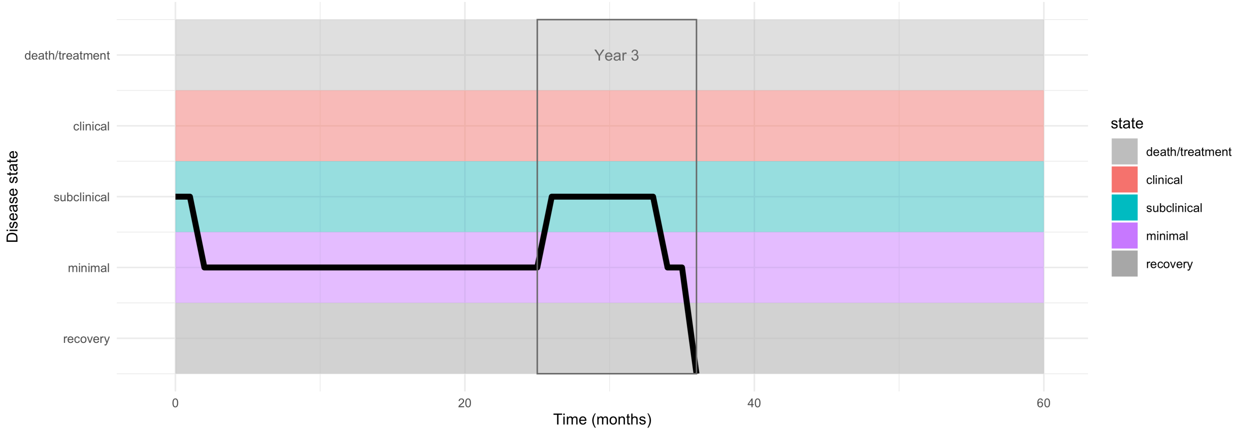


*Figure 10: An example trajectory where the individual recovers from TB. In this case, the individual spent the first 2 years having regressed to minimal, then progresses to subclinical in the third year before regressing quickly to recovery*

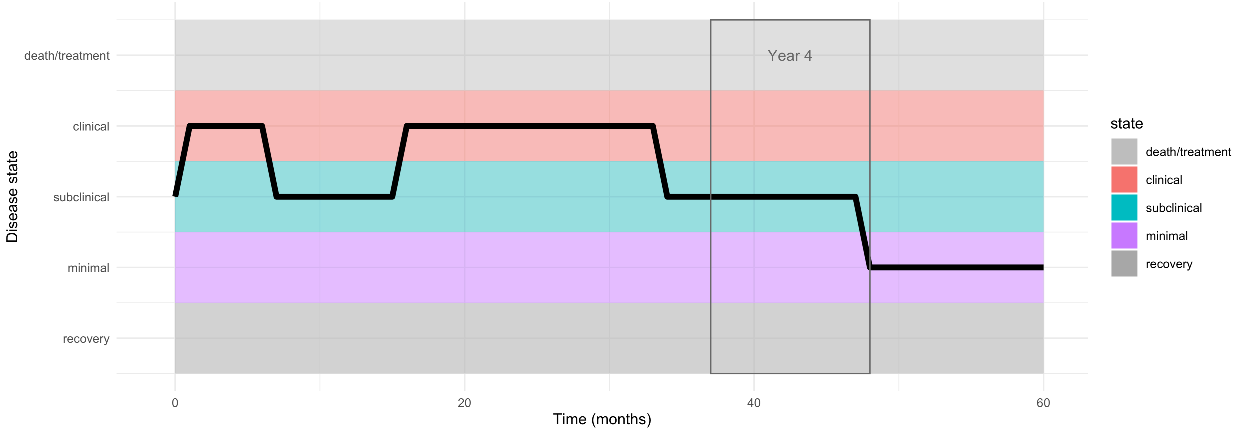


*Figure 11: An example trajectory of subclinical in the 4th year. The previous years have time in both clinical and subclinical and year 5 is entirely in minimal, however, as the majority of time (>=9 months) in year 4 and there are fewer than three state changes, the 4th year is defined as subclinical*

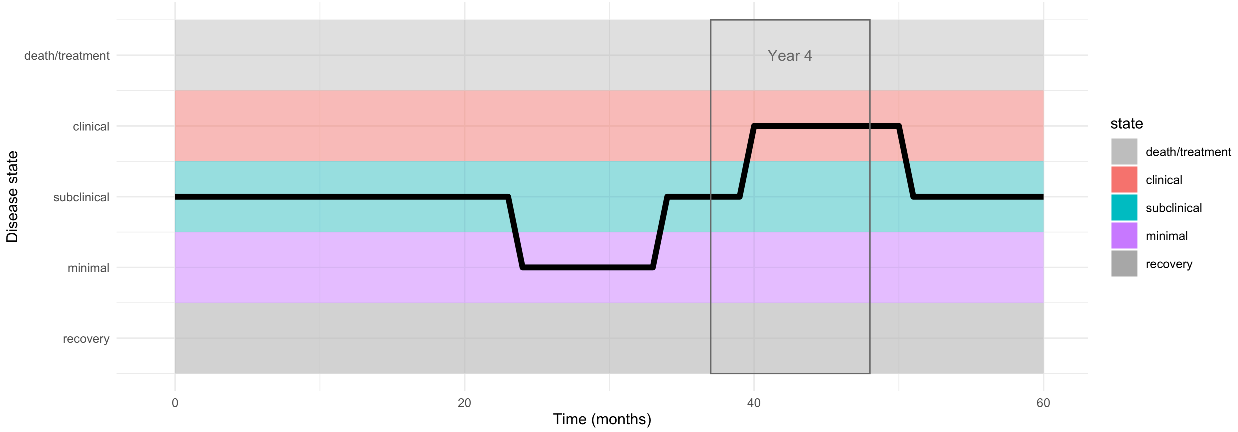


*Figure 12: An example trajectory of clinical in the 4th year. The previous years have time in both minimal and subclinical and year 5 is mainly subclinical. As the majority of time (>=9 months) in year 4 is clinical and there are fewer than three state changes, the 4th year is defined as clinical*

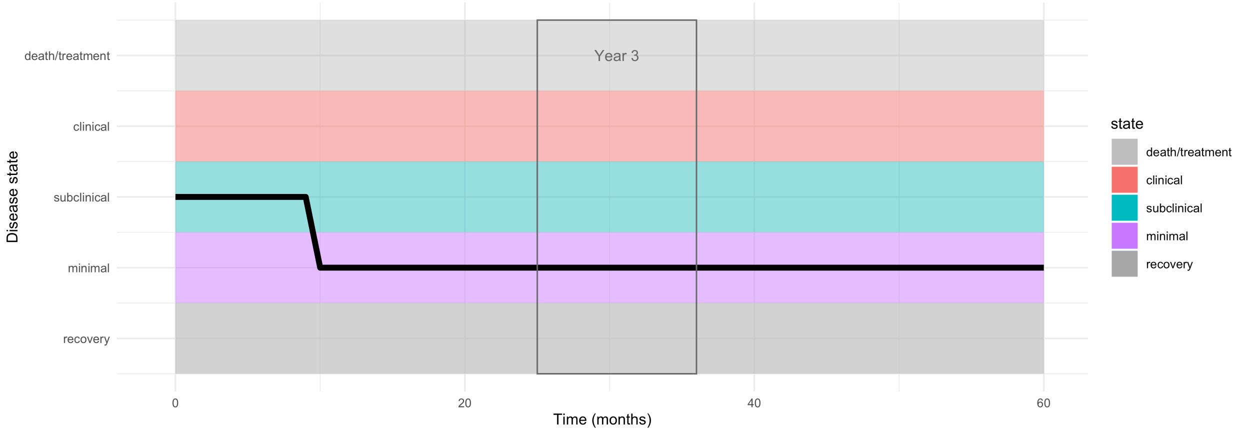


*Figure 13: An example trajectory of minimal in the 3rd year. Other than the first year, with time in subclinical, the remaining years are also all minimal*

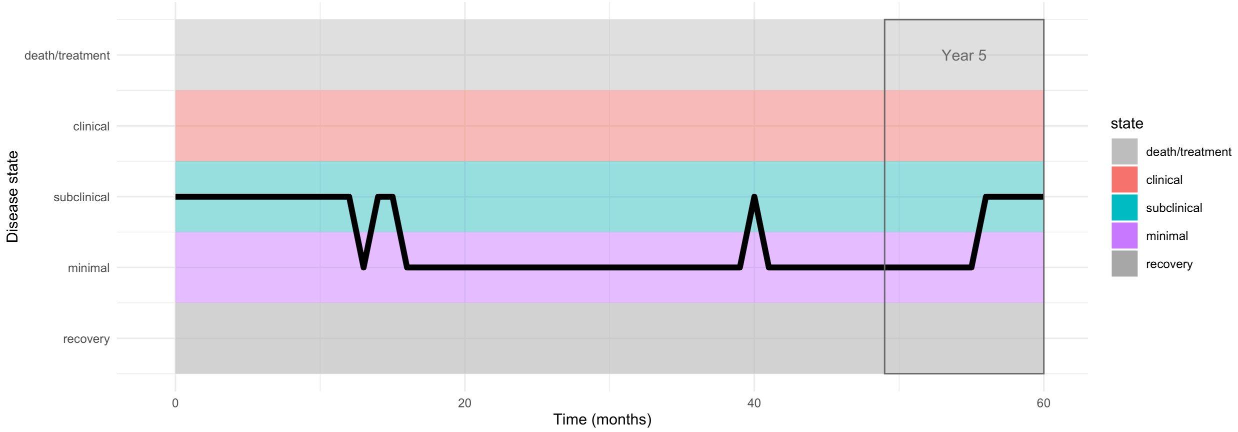


*Figure 14: An example trajectory of undulating disease in the 5th year. There are 2 disease states in the 5th year, with neither lasting for 9 months. The majority of the remainder of this trajectory is in minimal*

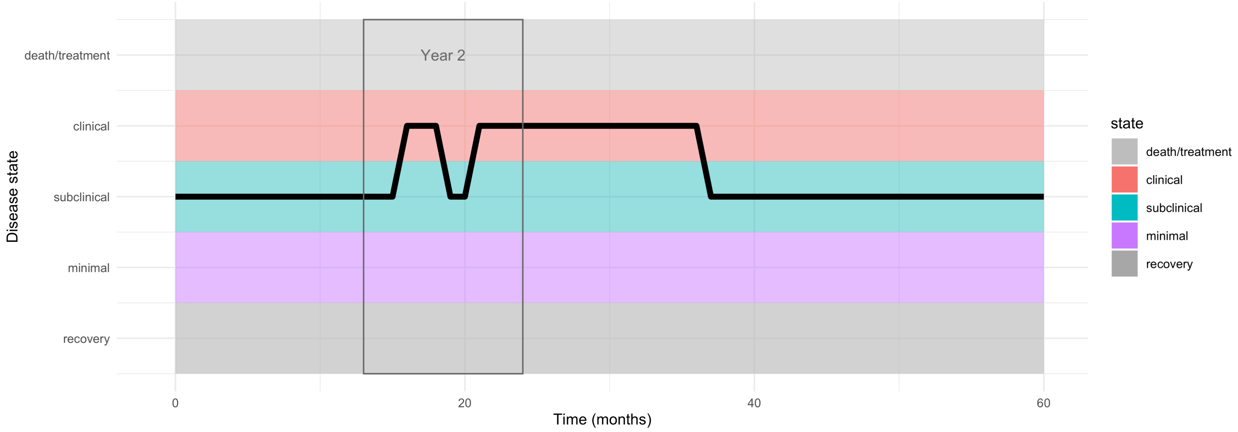


*Figure 15: An example trajectory of undulating disease in the second year, with three state changes and neither state lasting for a total of 9 months.*

### S8 Duration of symptoms

Duration of symptoms can be split into three categories; duration before death, duration before regression to subclinical, and when applicable, duration before treatment. These three have not shown a significant difference in our analysis, but of note is the highly skewed distribution that we observe. Of those who become clinical, the minimum time spent clinical is one month (as that is the time step in the model), but a small proportion of individuals have persistant symptoms for a long time

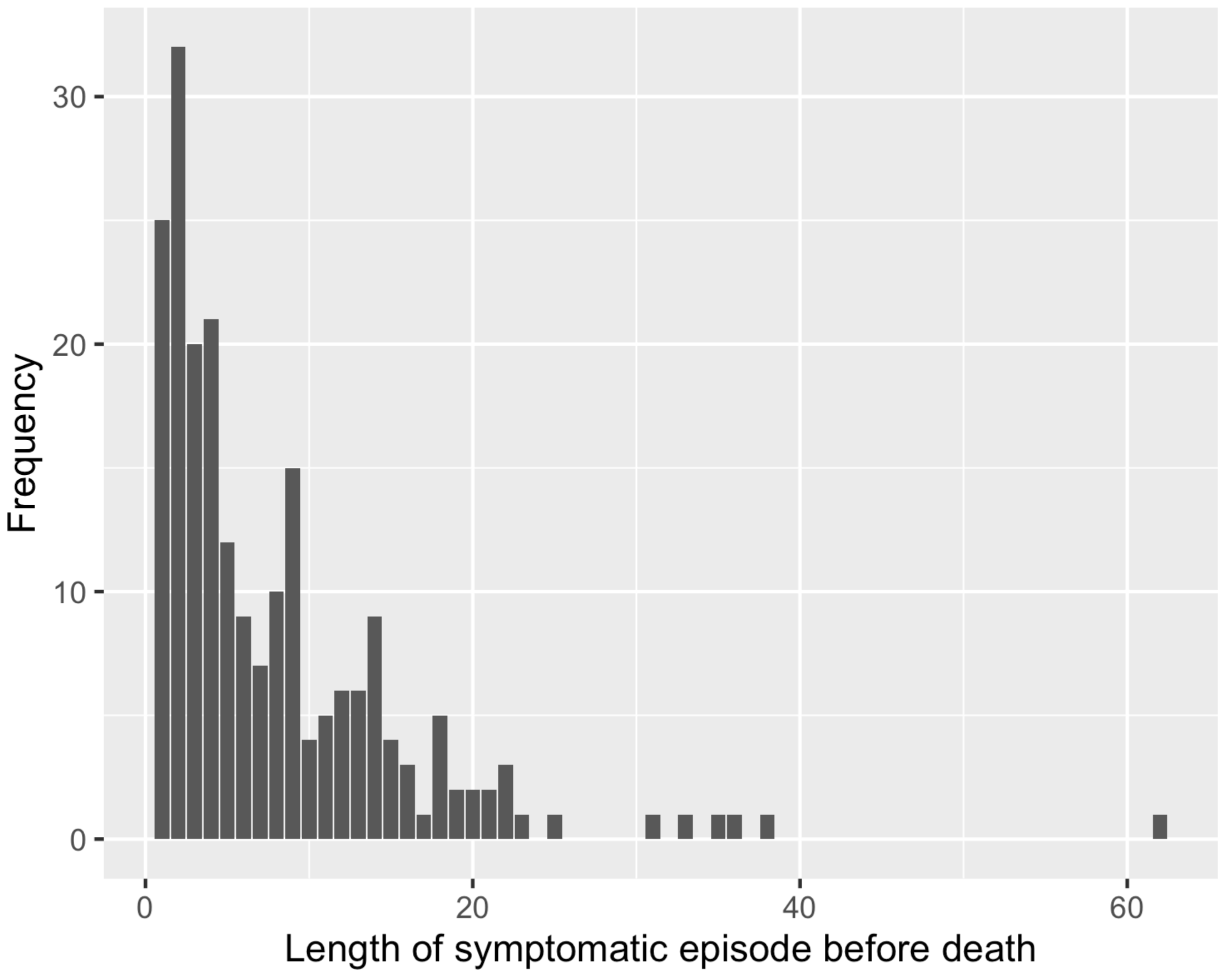
30
20
10
0
Frequency
0
20
40
60
Length of symptomatic episode before death

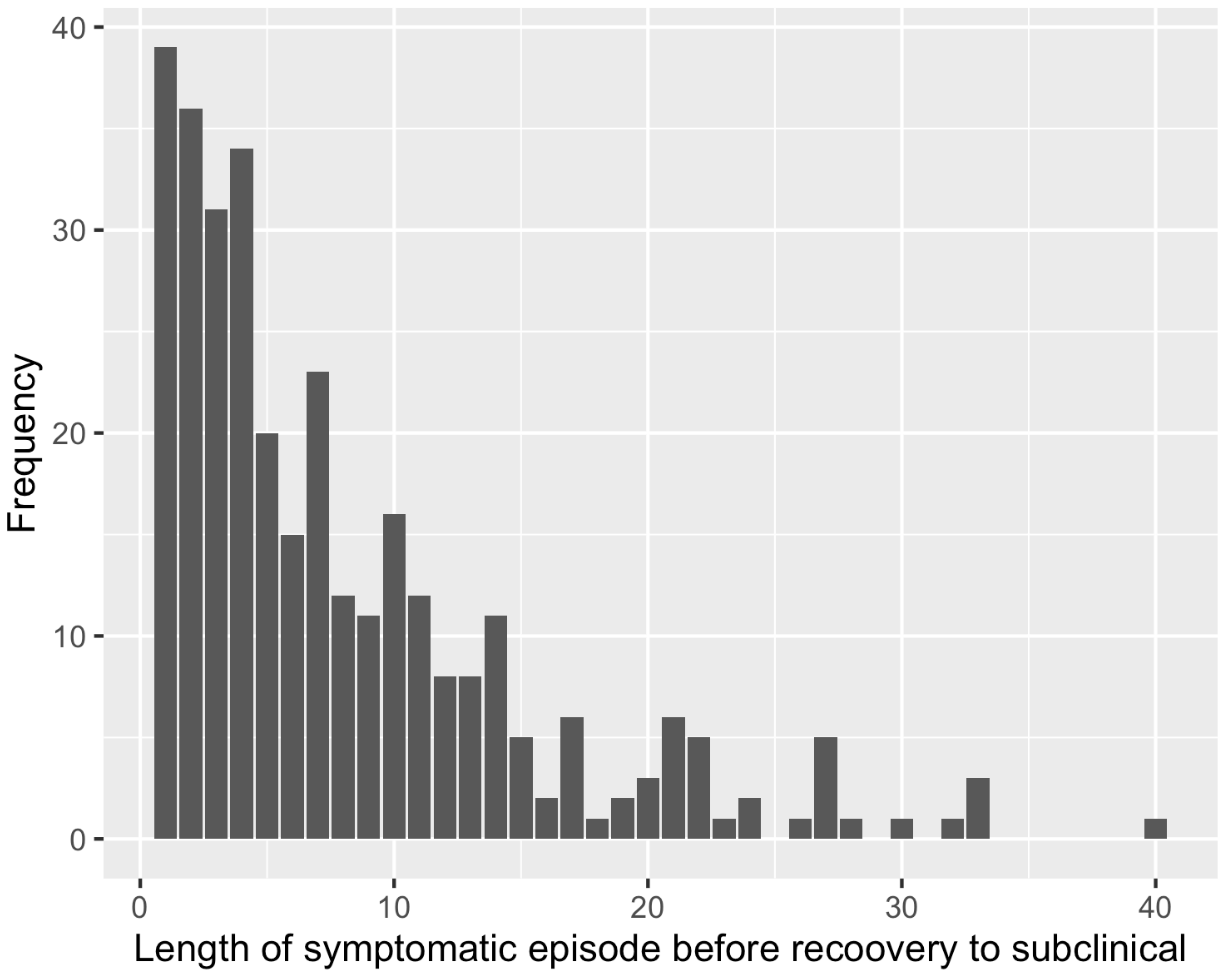

40
30
20
10
0
Frequency
0
10
20
30
40
Length of symptomatic episode before recoovery to subclinical

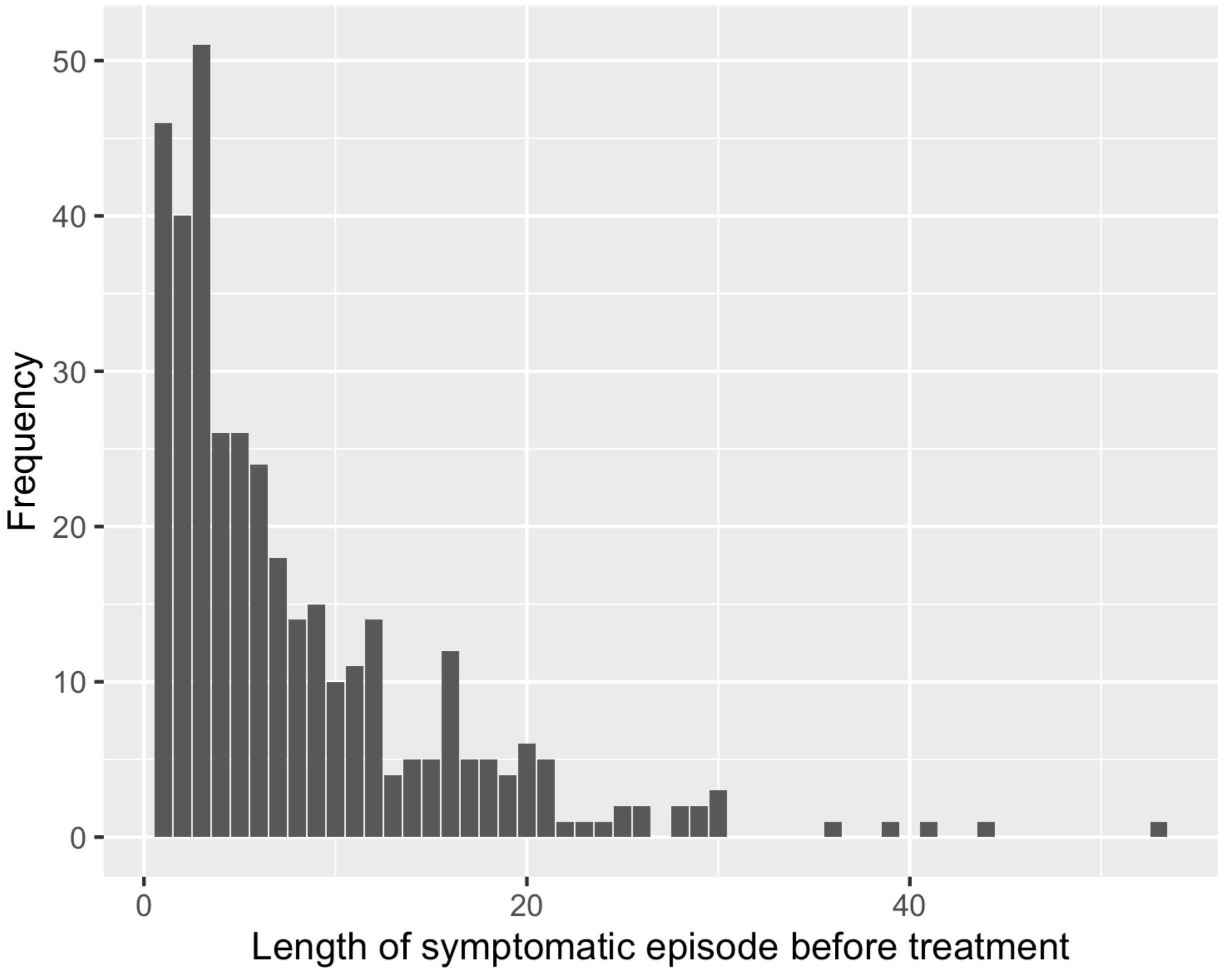


### S9 Additional results

Here we consider the median duration of disease and the proportion of people in each state at a given time. We can see that including treatment decreases the duration of disease and decreases the proportion clinical. When including minimal disease in the duration, we see that duration increases significantly, showing the importance of considering all those who are at risk of progressing to infectious disease. In the following figures, the top row is the number of people in all disease states (minimal, subclinical and clinical) over time, and the bottom row is the number of people in infectious disease states (subclinical and clinical).

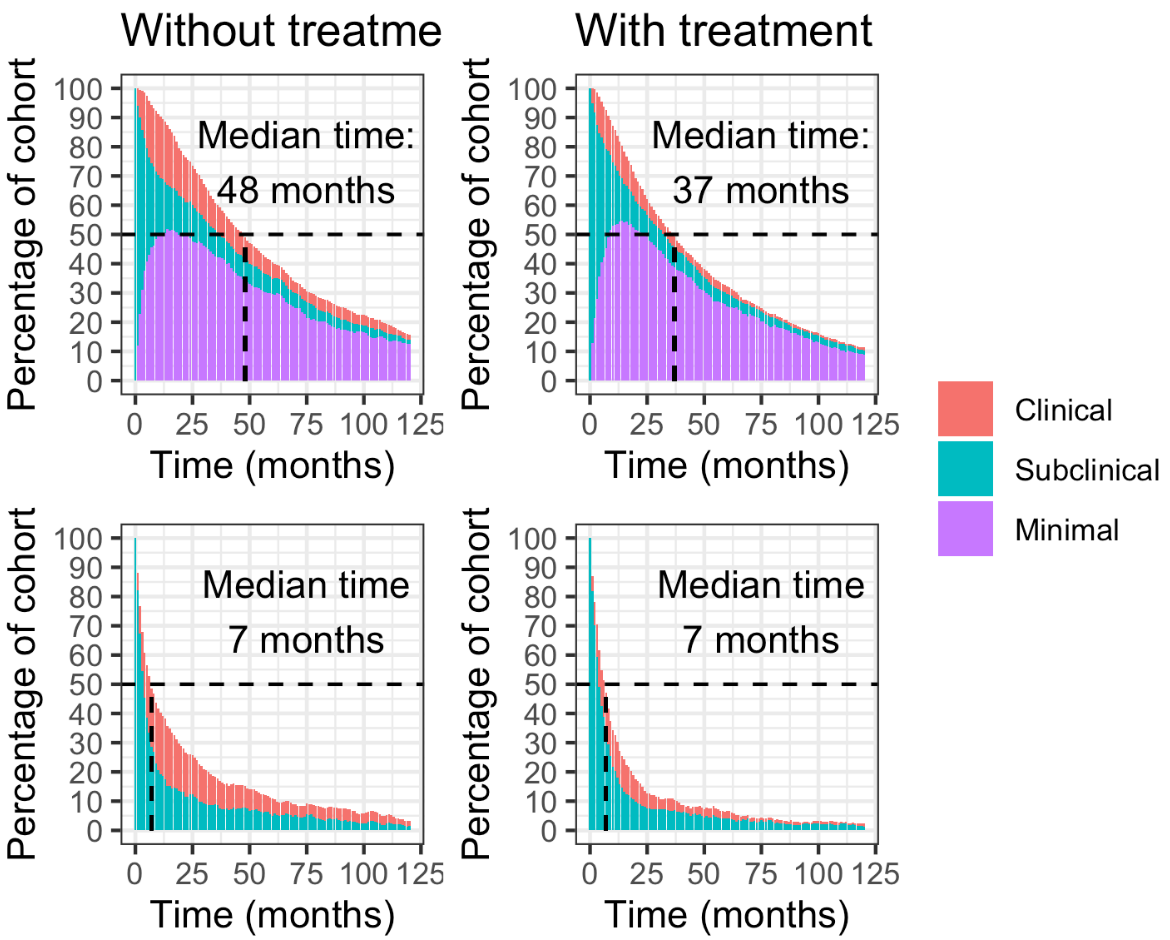


*Figure 16: Median duration of infectious and all disease with and without treatment, starting with a cohort of subclinical individuals*

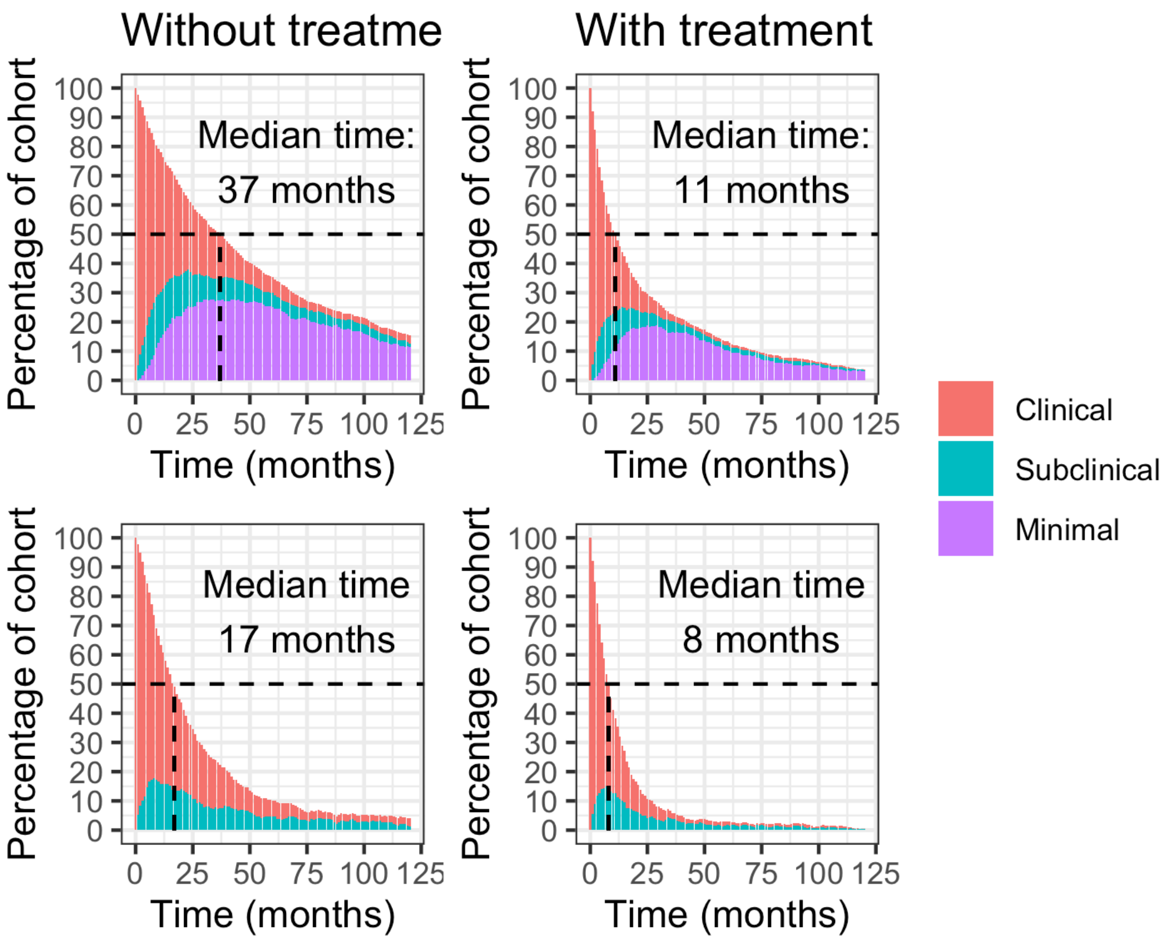


*Figure 17: Median duration of infectious and all disease with and without treatment, starting with a cohort of clinical individuals*

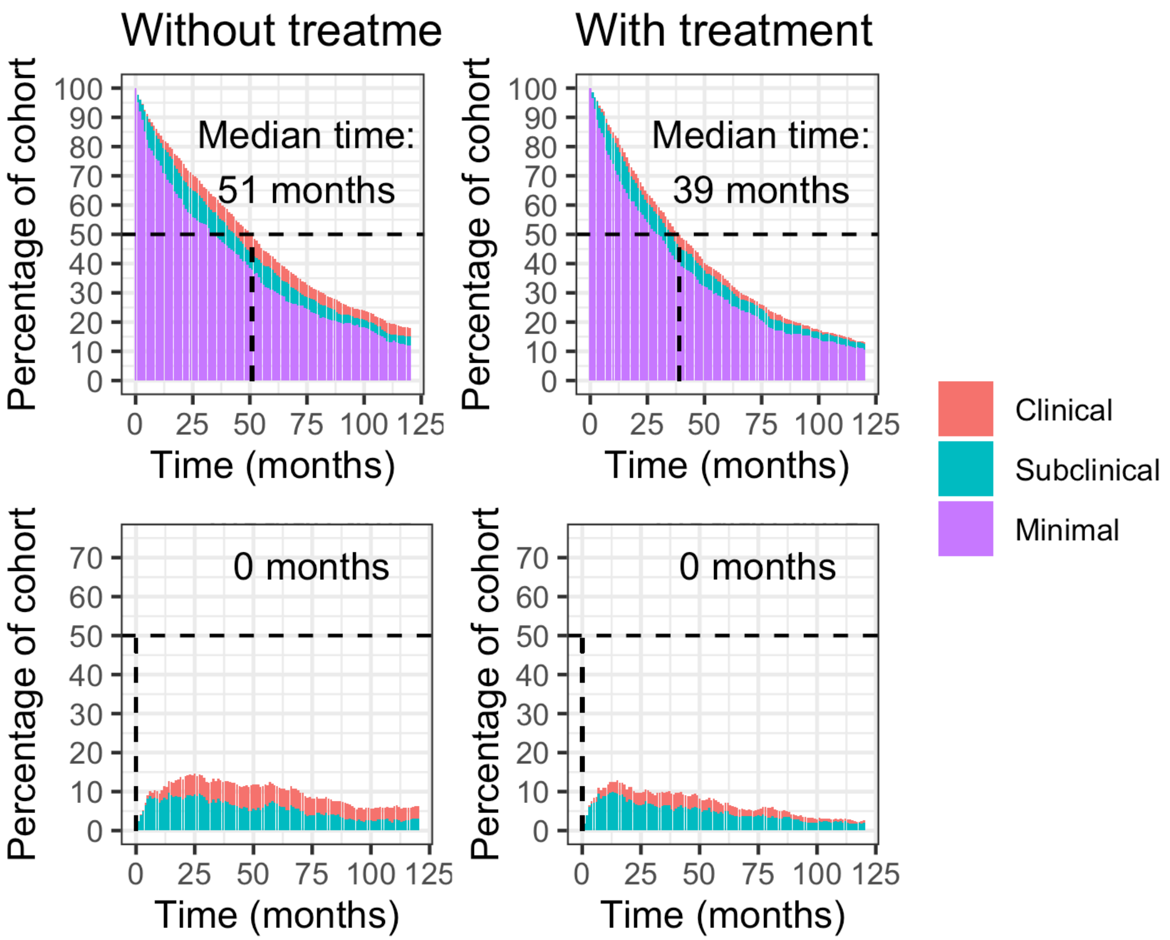


*Figure 18: Median duration of infectious and all disease with and without treatment, starting with a cohort of minimal individuals*

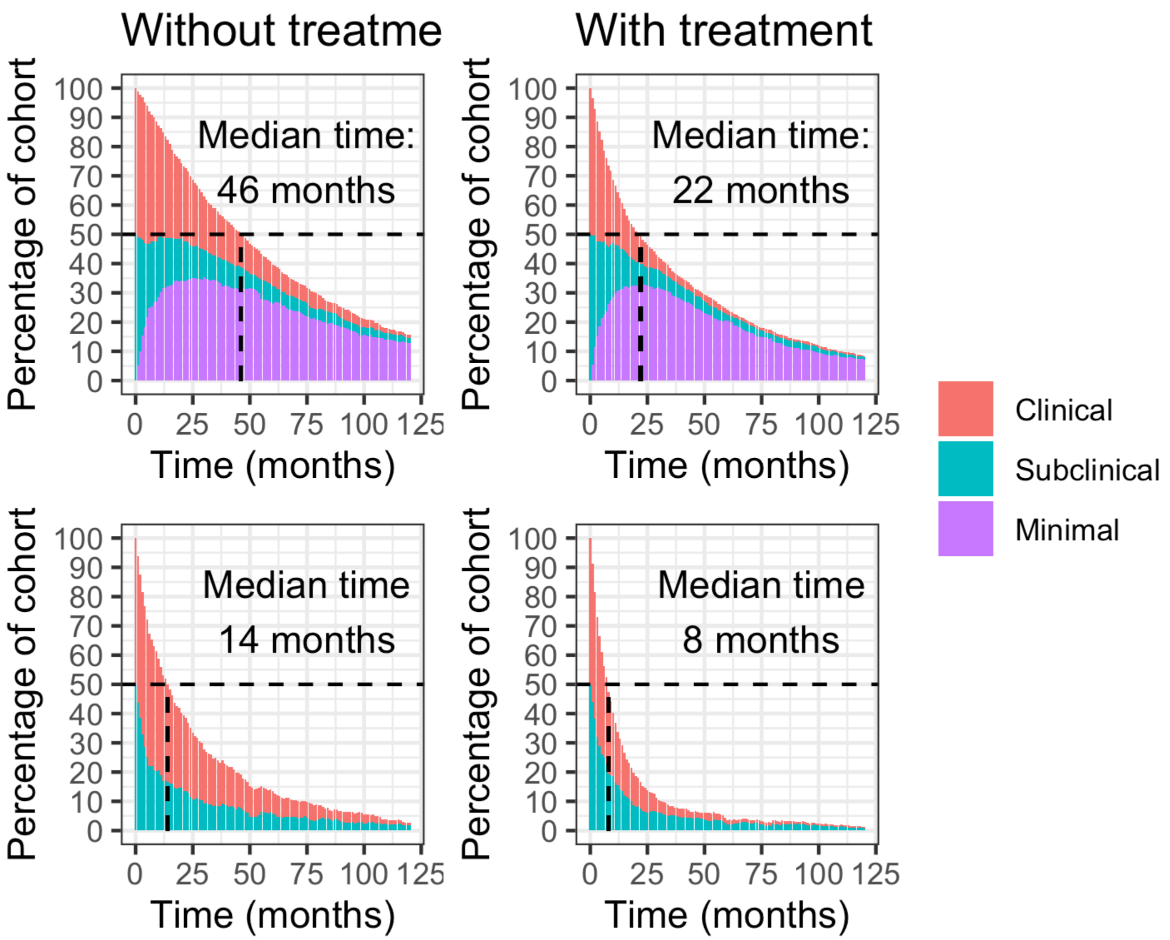


*Figure 19: Median duration of infectious and all disease with and without treatment, starting with a cohort of half clinical and half subclinical individuals*

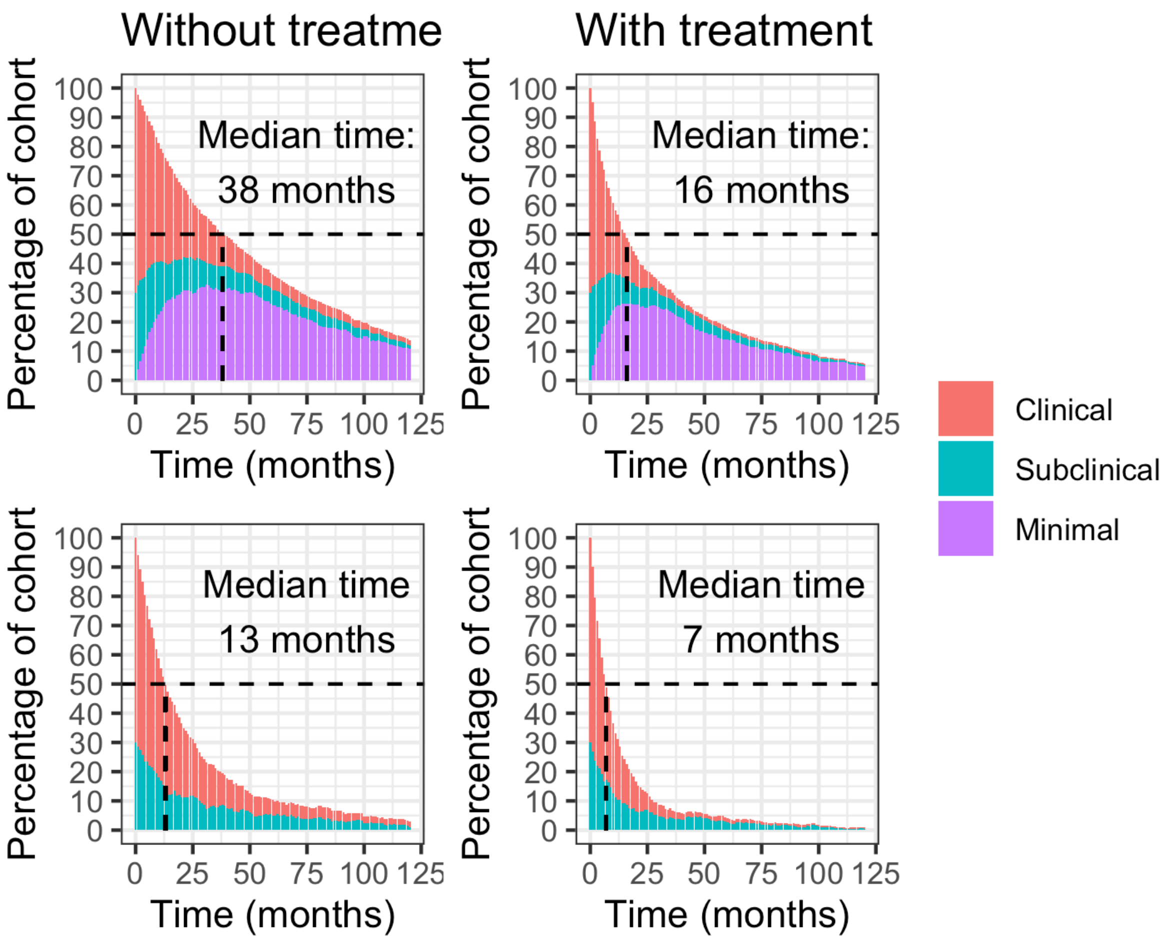


*Figure 20: Median duration of infectious and all disease with and without treatment, starting with a cohort of 70% clinical and 30% subclinical individuals*

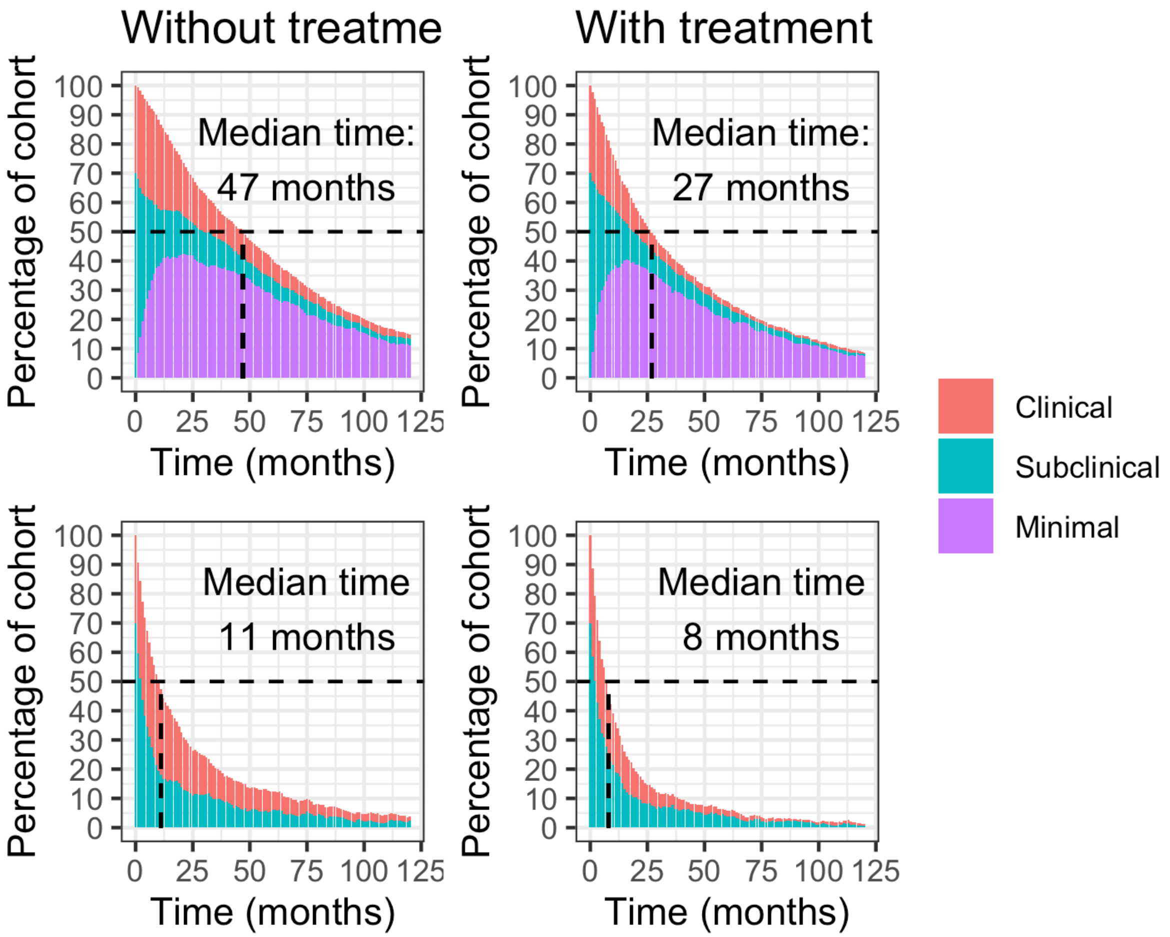


*Figure 21: Median duration of infectious and all disease with and without treatment, starting with a cohort of 30% clinical and 70% subclinical individuals*

### S10 Sensitivity Analyses

We have run sensitivity analyses on different areas of the main analysis. For the purposes of comparison, we have included the median parameter estimates, and key outputs; median duration of disease, percentage of people clinical, undulating, subclinical, and minimal after 5 years, and the number of people who have died from TB over 10 years.

#### S10.1 Fitting

##### *S10.1.1Bootstrap studies*

We ran the fit process removing the data from one study at a time. Table 4 shows the key outputs, showing that no one study is driving the fit, with similar results when each study is removed.

*Table 4: A summary if the differences in fit when removing one study at a time*

| study | min-out | min-sub | sub-min | sub-clin | clin-sub | mort | duration | clinical5 | undulating5 | subclinical5 | minimal5 | dead10 | treated5 | recovered5 |
|---|---|---|---|---|---|---|---|---|---|---|---|---|---|---|
| Main | 0.19 | 0.26 | 1.54 | 0.67 | 0.57 | 0.32 | 12 | 4.87 | 6.70 | 2.30 | 24.65 | 37.54 | 0 | 29.57 |
| 39-42 | 0.21 | 0.14 | 0.70 | 1.13 | 1.22 | 0.32 | 19 | 4.36 | 8.78 | 4.11 | 24.29 | 40.31 | 0 | 24.82 |
| #6897 | 0.20 | 0.14 | 0.69 | 1.23 | 1.30 | 0.32 | 18 | 4.39 | 8.76 | 4.09 | 22.79 | 39.86 | 0 | 26.60 |
| 12 | 0.20 | 0.14 | 0.71 | 1.12 | 1.17 | 0.32 | 18 | 4.50 | 8.48 | 4.03 | 23.45 | 39.87 | 0 | 26.04 |
| 10 | 0.21 | 0.14 | 0.65 | 1.07 | 1.26 | 0.33 | 18 | 3.88 | 8.93 | 4.42 | 22.44 | 39.64 | 0 | 26.77 |
| 28-36 | 0.23 | 0.14 | 0.66 | 1.27 | 1.35 | 0.33 | 18 | 4.22 | 9.20 | 3.93 | 22.15 | 38.61 | 0 | 28.12 |
| 38 | 0.19 | 0.13 | 0.65 | 0.88 | 1.08 | 0.33 | 19 | 4.60 | 8.05 | 5.07 | 24.81 | 37.78 | 0 | 26.00 |
| 13,14 | 0.22 | 0.14 | 0.74 | 1.16 | 1.21 | 0.33 | 17 | 4.46 | 8.37 | 3.83 | 22.73 | 38.58 | 0 | 28.24 |
| 20,21 | 0.21 | 0.14 | 0.70 | 1.11 | 1.17 | 0.32 | 18 | 4.15 | 8.39 | 4.05 | 23.00 | 39.75 | 0 | 26.69 |
| 45 | 0.20 | 0.14 | 0.70 | 1.16 | 1.24 | 0.33 | 18 | 4.03 | 8.62 | 3.99 | 22.94 | 40.83 | 0 | 26.28 |
| 43 | 0.21 | 0.14 | 0.69 | 1.16 | 1.24 | 0.32 | 18 | 4.02 | 9.16 | 3.87 | 23.77 | 39.55 | 0 | 26.11 |
| 11 | 0.23 | 0.14 | 0.69 | 1.16 | 1.25 | 0.33 | 19 | 3.94 | 8.87 | 4.12 | 21.56 | 39.80 | 0 | 27.93 |
| 18 | 0.20 | 0.14 | 0.68 | 1.21 | 1.30 | 0.33 | 18 | 4.11 | 9.15 | 4.31 | 23.20 | 39.99 | 0 | 25.90 |
| 44 | 0.21 | 0.18 | 0.95 | 0.88 | 0.83 | 0.32 | 15 | 5.08 | 7.32 | 3.34 | 23.36 | 38.47 | 0 | 28.58 |

| study | min-out | min-sub | sub-min | sub-clin | clin-sub | mort | duration | clinical5 | undulating5 | subclinical5 | minimal5 | dead10 | treated5 | recovered5 |
|---|---|---|---|---|---|---|---|---|---|---|---|---|---|---|
| 9 | 0.22 | 0.14 | 0.73 | 1.33 | 1.51 | 0.32 | 17 | 3.05 | 9.13 | 3.54 | 23.48 | 36.75 | 0 | 29.41 |
| 37 | 0.18 | 0.12 | 0.57 | 2.17 | 2.38 | 0.33 | 20 | 2.53 | 13.77 | 3.00 | 23.45 | 42.89 | 0 | 22.39 |
| 8 | 0.21 | 0.13 | 0.70 | 1.38 | 1.34 | 0.32 | 18 | 4.31 | 9.33 | 3.74 | 22.34 | 41.13 | 0 | 25.89 |
| 24 | 0.20 | 0.14 | 0.67 | 1.14 | 1.24 | 0.33 | 18 | 4.27 | 9.00 | 4.40 | 22.93 | 39.22 | 0 | 26.49 |
| 23 | 0.21 | 0.16 | 0.70 | 0.96 | 1.08 | 0.32 | 19 | 4.64 | 9.53 | 4.88 | 22.26 | 38.91 | 0 | 26.62 |
| 22 | 0.20 | 0.14 | 0.69 | 0.94 | 0.97 | 0.31 | 19 | 5.00 | 8.66 | 4.64 | 23.16 | 39.73 | 0 | 25.45 |
| 16,17 | 0.19 | 0.14 | 0.66 | 1.33 | 1.36 | 0.34 | 18 | 4.19 | 9.15 | 3.81 | 23.98 | 41.80 | 0 | 23.97 |
| 16,19 | 0.20 | 0.14 | 0.67 | 1.20 | 1.30 | 0.32 | 18 | 3.88 | 9.49 | 4.09 | 23.95 | 39.24 | 0 | 25.96 |
| 27 | 0.21 | 0.14 | 0.71 | 1.08 | 1.16 | 0.33 | 18 | 4.49 | 8.52 | 4.13 | 23.27 | 39.44 | 0 | 26.65 |

#### *S10.1.2Change duration of infectiousness*

The duration of infectiousness holds a fixed value of 2 years in the main analysis. In table 5 we change this fixed value to 18 months and 3 years.

*Table 5: A summary of the differences in fit when testing different durations of infectiousness*

| duration | min-out | min-sub | sub-min | sub-clin | clin-sub | mort | duration | clinical5 | undulating5 | subclinical5 | minimal5 | dead10 | treated5 | recovered5 |
|---|---|---|---|---|---|---|---|---|---|---|---|---|---|---|
| Main | 0.19 | 0.26 | 1.54 | 0.67 | 0.57 | 0.32 | 12 | 4.87 | 6.70 | 2.30 | 24.65 | 37.54 | 0 | 29.57 |

| duration | min-out | min-sub | sub-min | sub-clin | clin-sub | mort | duration | clinical5 | undulating5 | subclinical5 | minimal5 | dead10 | treated5 | recovered5 |
|---|---|---|---|---|---|---|---|---|---|---|---|---|---|---|
| 18 months | 0.21 | 0.14 | 0.73 | 1.06 | 1.11 | 0.32 | 18 | 4.19 | 8.28 | 4.09 | 24.07 | 38.96 | 0 | 26.76 |
| 3 years | 0.20 | 0.13 | 0.63 | 1.24 | 1.39 | 0.32 | 19 | 4.34 | 9.09 | 4.69 | 23.69 | 40.17 | 0 | 24.72 |

#### *S10.1.3Change proportion of minimal "true minimals"*

*Table 6: A summary of the differences in fit when changing the proportion of people that have x-ray changes due to TB disease*

| prop_min | min-out | min-sub | sub-min | sub-clin | clin-sub | mort | duration | clinical5 | undulating5 | subclinical5 | minimal5 | dead10 | treated5 | recovered5 |
|---|---|---|---|---|---|---|---|---|---|---|---|---|---|---|
| Main | 0.19 | 0.26 | 1.54 | 0.67 | 0.57 | 0.32 | 12 | 4.87 | 6.70 | 2.30 | 24.65 | 37.54 | 0 | 29.57 |
| 100% | 0.25 | 0.11 | 0.61 | 1.16 | 1.37 | 0.31 | 19 | 4.18 | 8.96 | 4.57 | 20.71 | 37.66 | 0 | 29.72 |
| 60% | 0.25 | 0.11 | 0.63 | 1.08 | 1.22 | 0.30 | 19 | 4.40 | 8.81 | 4.63 | 21.27 | 36.12 | 0 | 30.33 |

#### *S10.1.4Change assummption on persistent symptoms*

The studies where there was only information about the symptom state at the start, in the main analysis, we have assumed this symptom state persists. These data were from minimal to either subclinical or clinical, and so for this sensitivity, we have changed them all to minimal to infectious. The results are in table 7.

*Table 7: A summary of the difference in fit, and the subsequent analyses, when removing the assumption that symptoms persist*

| symptoms assumption | min-out | min-sub | sub-min | sub-clin | clin-sub | mort | duration | clinical5 | undulating5 | subclinical5 | minimal5 | dead10 | treated5 | recovered5 |
|---|---|---|---|---|---|---|---|---|---|---|---|---|---|---|
| Main | 0.19 | 0.26 | 1.54 | 0.67 | 0.57 | 0.32 | 12 | 4.87 | 6.70 | 2.30 | 24.65 | 37.54 | 0 | 29.57 |
| no | 0.20 | 0.17 | 0.85 | 0.77 | 0.81 | 0.32 | 16 | 4.66 | 7.18 | 4.14 | 23.75 | 38.04 | 0 | 27.90 |

### S10.2 Cohort model

#### *S10.2.1Parameter values*

The method of parameter choice for the simulation was one of three. In the main analysis, each step of the model, for each individual, the relevant parameters were chosen randomly from the posterior distribution. For the other two alternatives, we randomly sampled the parameters at the start of the simulation for each individual and fixed them for the whole run, and the other used the median parameters for each person. Table 8 shows that there is very little difference between either method overall.

*Table 8: A summary of the differences in analyis when testing different method of parameter choice for the cohort model*

| method | min-out | min-sub | sub-min | sub-clin | clin-sub | mort | duration | clinical5 | undulating5 | subclinical5 | minimal5 | dead10 | treated5 | recovered5 |
|---|---|---|---|---|---|---|---|---|---|---|---|---|---|---|
| Main | 0.19 | 0.26 | 1.54 | 0.67 | 0.57 | 0.32 | 12 | 4.87 | 6.70 | 2.30 | 24.65 | 37.54 | 0 | 29.57 |
| fixed | 0.19 | 0.26 | 1.54 | 0.67 | 0.57 | 0.32 | 12 | 4.75 | 7.20 | 2.03 | 24.67 | 37.35 | 0 | 29.37 |
| median | 0.19 | 0.26 | 1.54 | 0.67 | 0.57 | 0.32 | 12 | 4.58 | 6.63 | 2.14 | 24.16 | 38.74 | 0 | 29.64 |

#### *S10.2.2Treatment*

Treatment was added to the model to simulate a case detection rate for a care pathway initiated by self reported symptoms. When considered in the main analysis, the case detection rate was implemented at 70%. In table 9 we compare the difference between case detection rates at 50%, 70%, and 90%.

*Table 9: A summary of the differences in analysis when testing different passive case detection rates*

| treatment | min-out | min-sub | sub-min | sub-clin | clin-sub | mort | duration | clinical5 | undulating5 | subclinical5 | minimal5 | dead10 | treated5 | recovered5 |
|---|---|---|---|---|---|---|---|---|---|---|---|---|---|---|
| Main | 0.19 | 0.26 | 1.54 | 0.67 | 0.57 | 0.32 | 12 | 4.87 | 6.70 | 2.30 | 24.65 | 37.54 | 0.00 | 29.57 |
| 0.5 | 0.19 | 0.26 | 1.54 | 0.67 | 0.57 | 0.32 | 8 | 1.21 | 4.06 | 1.24 | 18.44 | 21.76 | 29.16 | 26.11 |
| 0.7 | 0.19 | 0.26 | 1.54 | 0.67 | 0.57 | 0.32 | 7 | 0.79 | 3.64 | 1.01 | 17.40 | 19.03 | 33.89 | 25.54 |

| treatment | min-out | min-sub | sub-min | sub-clin | clin-sub | mort | duration | clinical5 | undulating5 | subclinical5 | minimal5 | dead10 | treated5 | recovered5 |
|---|---|---|---|---|---|---|---|---|---|---|---|---|---|---|
| 0.9 | 0.19 | 0.26 | 1.54 | 0.67 | 0.57 | 0.32 | 7 | 0.56 | 2.91 | 1.11 | 16.77 | 16.65 | 39.32 | 23.81 |

#### *S10.2.3Trajectories*

The trajectories are based on two variables, the proportion of time in a single state, and the number of state changes both over the previous 12 months. The main analysis defines undulating as less than nine months in a single state or 3 or more changes in state. In table 10 we compare the definition of undulation as less than 8 months or less than 10 months, whilst keeping the number of state changes fixed at 3 or more. In table 11 we compare the definition of undulation with less than 9 months fixed, and the state changes as either 2 or more, or 4 or more.

*Table 10: A summary of the differences in analysis when varying the threshold for undulating trajectories*

| states | min-out | min-sub | sub-min | sub-clin | clin-sub | mort | duration | clinical5 | undulating5 | subclinical5 | minimal5 | dead10 | treated5 | recovered5 |
|---|---|---|---|---|---|---|---|---|---|---|---|---|---|---|
| Main | 0.19 | 0.26 | 1.54 | 0.67 | 0.57 | 0.32 | 12 | 4.87 | 6.70 | 2.30 | 24.65 | 37.54 | 0 | 29.57 |
| 7 | 0.19 | 0.26 | 1.54 | 0.67 | 0.57 | 0.32 | 12 | 5.48 | 2.67 | 4.02 | 25.84 | 37.64 | 0 | 30.23 |
| 8 | 0.19 | 0.26 | 1.54 | 0.67 | 0.57 | 0.32 | 12 | 5.32 | 4.61 | 2.75 | 25.36 | 37.54 | 0 | 30.26 |
| 10 | 0.19 | 0.26 | 1.54 | 0.67 | 0.57 | 0.32 | 12 | 4.15 | 9.17 | 1.52 | 23.28 | 37.52 | 0 | 30.01 |
| 11 | 0.19 | 0.26 | 1.54 | 0.67 | 0.57 | 0.32 | 12 | 3.89 | 11.76 | 1.05 | 21.86 | 37.82 | 0 | 29.50 |

*Table 11: A summary of the differences in analysis when varying the threshold for undulating trajectories*

| changes | min-out | min-sub | sub-min | sub-clin | clin-sub | mort | duration | clinical5 | undulating5 | subclinical5 | minimal5 | dead10 | treated5 | recovered5 |
|---|---|---|---|---|---|---|---|---|---|---|---|---|---|---|
| Main | 0.19 | 0.26 | 1.54 | 0.67 | 0.57 | 0.32 | 12 | 4.87 | 6.70 | 2.30 | 24.65 | 37.54 | 0 | 29.57 |
| 2 | 0.19 | 0.26 | 1.54 | 0.67 | 0.57 | 0.32 | 12 | 3.92 | 9.47 | 1.71 | 23.22 | 37.68 | 0 | 29.13 |
| 4 | 0.19 | 0.26 | 1.54 | 0.67 | 0.57 | 0.32 | 12 | 5.11 | 6.55 | 2.09 | 24.19 | 37.61 | 0 | 29.93 |

### References


1 Tiemersma EW, Werf MJ van der, Borgdorff MW, Williams BG, Nagelkerke NJD. Natural history of tuberculosis: Duration and fatality of untreated pulmonary tuberculosis in HIV negative patients: A systematic review. *PLoS ONE* 2011; **6**: e17601.

2 Ragonnet R, Flegg JA, Brilleman SL, Tiemersma EW, Melsew YA, McBryde ES *et al.* Revisiting the natural history of pulmonary tuberculosis: A bayesian estimation of natural recovery and mortality rates. *Clin Infect Dis* 2020. doi:10.1093/cid/ciaa602.

3 Szucs E. Sputum examination in so-called closed tuberculosis. *JAMA* 1926; **86**: 946.

4 Tattersall WH. The survival of sputum-positive consumptives: A study of 1,192 cases in a county borough between 1914 and 1940. *Tubercle* 1947; **28**: 85–96.

5 Lindhardt M. *The statistics of pulmonary tuberculosis in denmark, 1925-1934; a statistical investigation on the occurrence of pulmonary tuberculosis in the period 1925-1934, worked out on the basis of the danish national health service file of notified cases and of deaths.* EMunksgaard: Copenhagen, 1939.

6 Blahd M, Leslie EI, Rosenthal SR. Infectiousness of the 'closed case' in tuberculosis. *Am J Public Health Nations Health* 1946; **36**: 723–726.

7 Association NT. *Diagnostic standards and classification of tuberculosis.* 1940 ed. New York, N.Y., 1940http://hdl.handle.net/2027/coo.31924089435949.

8 Okada K, Onozaki I, Yamada N, Yoshiyama T, Miura T, Saint S *et al.* Epidemiological impact of mass tuberculosis screening: A 2 year follow-up after a national prevalence survey. 2012; **NA**.

9 Downes J. The study of mortality among individuals with active pulmonary tuberculosis. 1938; **16**: 304–317.

10 Beeuwkes H, Hahn RG, Putnam P. A survey of persons exposed to tuberculosis in the household. *Am Rev Tuberc* 1942; **45**: 165–193.

11 Puffer RR, Stewart HC, Gass RS. Tuberculosis according to age, sex, family history, and contact. *Am Rev Tuberc* 1945; **51**: 295–311.

12 Puelma HO, Grebe G. Analysis of one hundred cases of minimal pulmonary tuberculosis. *Dis Chest* 1945; **11**: 375–379.

13 BOBROWITZ ID., HURST A., MARTIN M. Minimal tuberculosis; the prognosis and clinical significance of a sanatorium treated group. *American review of tuberculosis* 1947; **56**: 110–125.

14 BOBROWITZ ID., HURST A. Minimal tuberculosis; problems in roentgenologic interpretation. *Radiology* 1949; **52**: 519–32.

15 Childress WG. Clinical evaluation, treatment, and follow-up of newly acquired tuberculous lesions. *New York State Journal of Medicine* 1947; **47**: 2560–4.

16 Bosworth E.B., Alling D.W. The after-history of pulmonary tuberculosis. I. Methods of evaluation. *American Review of Tuberculosis* 1954; **69**: 37–49.

17 Lincoln NS, Bosworth EB, Alling DW. The after-history of pulmonary tuberculosis. III. Minimal tuberculosis. *Am Rev Tuberc* 1954; **70**: 15–31.

18 Streptomycin treatment of pulmonary tuberculosis: A medical research council investigation. *BMJ* 1948; **2**: 769–782.

19 Alling D.W., Bosworth E.B., Lincoln N.S. The after-history of pulmonary tuberculosis. V. Moderately advanced tuberculosis. *American Review of Tuberculosis* 1955; **71**: 519–528.

20 Borgen L., Meyer SN., Refsum E. Mass photofluorography, tuberculin testing, and BCG vaccination in the district of aker (norway) 1947-49. *Acta tuberculosea Scandinavica* 1951; **25**: 327–55.

21 Refsum E. Mass investigation by photofluorography; an illustration of the value of the method in combating tuberculosis. *Acta tuberculosea Scandinavica* 1952; **27**: 288–302.

22 Manser H. [Tuberculosis in aged and its course during sanatorium treatment]. *Schweizerische Zeitschrift fur Tuberkulose Revue suisse de la tuberculose Rivista svizzera della tubercolosi* 1953; **10**: 65–82.

23 K. B. [Public health x-ray diagnosis of closed pulmonary tuberculosis later proved contagious]. *Beitrage zur Klinik der Tuberkulose und spezifischen Tuberkulose-Forschung* 1954; **111**: 437–44.

24 Sikand BK, Narain R, Mathur GP. Incidence of TB as judged by re-surveys. A study of delhi police. *Indian Journal of Tuberculosis* 1959; **6**: 73–83.

25 Tuberculosis Society of Scotland. A controlled trial of chemotherapy in pulmonary tuberculosis of doubtful activity. Report from the research committee of the tuberculosis society of scotland. *Tubercle* 1958; **39**: 129–137.

26 Society ST. A controlled trial of chemotherapy in pulmonary tuberculosis of doubtful activity. *Tubercle* 1963; **44**: 39–46.

27 Frimodt-Moller J. Results of treatment of non-bacillary tuberculosis in a domiciliary treatment programme, preliminary report. Proceedings of the 20th tuberculosis and chest diseases workers' conference. 1965, p 133.

28 National Tuberculosis Institute B. Tuberculosis in a rural population of south india: A five-year epidemiological study. *Bull World Health Organ* 1974; **51**: 473–88.

29 Chakraborty AK, Singh H, Srikantan K, Rangaswamy KR, Krishnamurthy MS, Stephen JA. Tuberculosis in a rural population of south india: Report on five surveys. *Indian Journal of Tuberculosis* 1982; **29**: 153–167.

30 Gothi GD, Chakraborty AK, Krishnamurthy VV, Banerjee GC. Prevalence and incidence of sputum negative active pulmonary tuberculosis and fate of pulmonary rradiological abnormalities found in rural population. *Indian Journal of Tuberculosis* 1978; **25**: 122–131.

31 Krishnamurthy VV, Nair SS, Gothi GD. A comparison of new cases (incidence cases) who had come from differeent epidemiological groups in the population. *Indian Journal of Tuberculosis* 1978; **25**: 144–146.

32 Gothi GD, Chakraborty AK, Jayalakshmi MJ. Incidence of sputum positive tuberculosis in different epidemiological grroups during five year follow up of a rural population in south india. *Indian Journal of Tuberculosis* 1978; **25**: 83–91.

33 Krishnamurthy VV, Nair SS, Gothi GD, Chakraborty AK. Incidence of tuberculosis among newly infected population and in relation to the duration of infected status. *Indian Journal of Tuberculosis* 1976; **23**.

34 Gothi GD, Nair SS, Chakraborty AK, Ganapathy KT. Five year incidencee of tuberculosis and crude mortality in relation to non-specific tuberculin senssitivity. *Indian Journal of Tuberculosis* 1976; **23**.

35 Chakraborty AK, Gothi GD. Relapses among naturally cureed cases of pulmmonary tuberculosis. *Indian Journal of Tuberculosis* 1976; **23**: 8–13.

36 Gothi GD, Chakraborty AK, Banerjee GC. Interpretation of photofluorograms of active pulmonary tuberculosis patients found in epidemiological survey and their five year fate. *Indian Journal of Tuberculosis* 1974; **11**: 90–97.

37 Pamra SP, Mathur GP. Effects of chemoprophylaxis on minimal pulmonary tuberculosis lesions of doubtful activity. *Bull World Health Organ* 1971; **45**: 593–602.

38 Aneja KS, Gothi GD, Samuel GER. Controlled study of the effect of spcific treatment on bacteriological status of ’suspect cases’. *Indian J Tuberc* 1979; **26**: 50–57.

39 Hong Kong Chest Service/Tuberculosis Research Centre MMRC. A controlled trial of 2-month, 3-month, and 12-month regimens of chemotherapy for sputum smear-negative pulmonary tuberculosis: The results at 30 months. *Am Rev Respir Dis* 1981; **124**: 138–142.

40 Hong Kong Chest Service/Tuberculosis Research Centre MMRC. A controlled trial of 2-month, 3-month, and 12-month regimens of chemotherapy for sputum-smear-negative pulmonary tuberculosis. *Am Rev Respir Dis* 1984; **130**: 23–28.

41 Hong Kong Chest Service/Tuberculosis Research Centre MMRC. Sputum smear negative pulmonary tuberculosis controlled trial of 3-month and 2-month regimend of chmotherapy: First report. *The Lancet* 1979; **313**: 1361–1363.

42 Hong Kong Chest Service/Tuberculosis Research Centre MMRC. A study of the characteristics and course of sputum smear-negative pulmonary tuberculosis. *Tubercle* 1981; **62**: 155–167.

43 Cowie RL. Diagnosis of sputum smear- and sputum culture-negative pulmonary tuberculosis. *SAMJ* 1985; **68**: 878.

44 Anastasatu C, Berceea O, Corlan E. Controlled clinical trial on smear negatice, x-ray positive new cases, with the view to establishing if and how to treat them. *Bull Int Union Tub* 1985; **60**: 108–109.

45 Norregaard J. Abacillary pulmonary tuberculosis. *Tubercle* 1990; **71**: 35–38.

46 Madsen T. Studies on the epidemiology of tuberculosis in denmark. *Acta Tuberculosea Scand (suppl)* 1942; **Supp 6**: 1–176.

47 Efficacy of various durations of isoniazid preventive therapy for tuberculosis: Five years of follow-up in the IUAT trial. *Bulletin of the World Health Organization* 1982; **60**: 555–564.

48 GROTH-PETERSEN E., KNUDSEN J., WILBEK E. Epidemiological basis of tuberculosis eradication in an advanced country. *Bulletin of the World Health Organization* 1959; **21**: 5–49.

49 Groth-Petersen E, Østergaard F. Mass chemoprophylaxis of tuberculosis. The acceptability and untoward side effects of isoniazid in a control study in greenland. *Am Rev Respir Dis* 1960; **81**: 643–652.

50 Styblo K, Dajkova D, Kubik A, Langerova M, Radkovsky J. Epidemiological and clinical study of tuberculosis in the district of kolin, czechoslovakia. ; : 56.

51 Rubinshteyn GR, Kochnova IE. Pulmonary tuberculosis: Inital stage and development in adults. *Acta Med,URSS* 1940; **3**: 250–265.

52 Frascella B, Richards AS, Sossen B, Emery JC, Odone A, Law I *et al.* Subclinical tuberculosis disease - a review and analysis of prevalence surveys to inform definitions, burden, associations and screening methodology. *Clin Infect Dis* 2020. doi:10.1093/cid/ciaa1402.
